# Moderate hypoxia and cognitive training for cognitive impairment in mood disorders: a randomized controlled trial

**DOI:** 10.64898/2026.07.10.26357722

**Authors:** Johanna Mariegaard Schandorff, Viktoria Damgaard, Annette Johansen, Julian Macoveanu, Katrine Cramer, Anne Bügel Fisker Madsen, Barbara Emilie Rotbøl Ørum, Caroline Fussing Bruun, Morten Meyer, Pontus Plavén-Sigray, Claus Svarer, Gitte Moos Knudsen, Martin Balslev Jørgensen, Lars Vedel Kessing, Hannelore Ehrenreich, Kamilla Woznica Miskowiak

**Affiliations:** The Neurocognition and Emotion Across Disorders of the Brain (NEAD) Centre, Psychiatric Centre Copenhagen, Frederiksberg, Denmark; Department of Psychology, University of Copenhagen, Copenhagen, Denmark; Neurobiology Research Unit, Copenhagen University Hospital, Copenhagen, Denmark; Department of Clinical Medicine, University of Copenhagen, Copenhagen, Denmark; Copenhagen Affective Disorders Research Center (CADIC), Psychiatric Centre Copenhagen, Frederiksberg, Denmark; Department of Neurobiology Research, Institute of Molecular Medicine, University of Southern Denmark, Odense, Denmark; Department of Neurology, Odense University Hospital, Odense, Denmark; BRIDGE – Brain Research Inter-Disciplinary Guided Excellence, Department of Clinical Research, University of Southern Denmark, Odense, Denmark; Department of Clinical Neuroscience, Centre for Psychiatry Research, Karolinska Institutet and Stockholm Health Care Services, Stockholm, Sweden; Experimental Medicine, Central Institute of Mental Health (CIMH), Mannheim, Germany

## Abstract

Cognitive impairment is a debilitating feature of mood disorders (MD) linked to decreased neuroplasticity. Moderate hypoxia upregulates neuroplasticity and may improve cognition when combined with cognitive training. In this outcome assessor-blinded randomized controlled trial, we investigate the effects of three weeks of repeated hypoxia (12% O_2_) with concurrent cognitive training (H-CT) for 3.5 hours, 5-6 days per week compared with treatment as usual (TAU). Cognitively impaired individuals with remitted MD were randomized to H-CT or TAU and assessed at baseline, treatment completion, and one-month follow-up. The primary outcome was a broad cognitive composite measure. Additional cognitive, functioning, blood-based, and neuroimaging (SV2A as a presynaptic marker with [^11^C]UCB-J positron emission tomography (PET) and neural activity during working memory functional magnetic resonance imaging (fMRI)) outcomes were assessed. Sixty-four participants were randomized to H-CT (*n*=34) or TAU (*n*=30), and 26 H-CT and 29 TAU completed the intervention and assessments. There was no effect of H-CT on the primary outcome (*ps≥*0.24) but H-CT induced improvements in the secondary executive function outcome (treatment effect=0.76, 95% CI=[0.23;1.28], adj. *p*=0.02) that prevailed at follow-up (treatment effect=1.07, 95% CI=[0.36;1.77], adj. *p*=0.02). Executive function improvement was accompanied by H-CT-related lower presynaptic [^11^C]UCB-J binding in the nucleus accumbens (*p*=0.03). H-CT also produced sustained improvements in everyday functioning (adj. *ps*≤0.049). No effects were observed on dorsolateral prefrontal cortex activity or real-life cognitive skills (*p*s≥0.54). Two serious but treatment-unrelated adverse events occurred. In conclusion, H-CT produces robust effects on executive functions, but not broader cognition, in MD, and H-CT may thus be particularly promising for targeting executive dysfunction in this population.

## Introduction

Mood disorders (MD) affect 320 million people worldwide and are among the top five of psychiatric conditions with greatest socio-economic costs [1], primarily driven by high unemployment rates and reduced productivity [2]. Cognitive impairments, including memory, concentration, and planning difficulties are among the key drivers of this functional disability [3]. When cognition is impaired, affected individuals struggle to work, complete their education and live independently. For approximately 50-70% of individuals with MD, cognitive impairments persist after symptomatic remission [4]. This has led to the recognition that cognitive impairments constitute a pressing treatment target to improve levels of functioning and reduce societal costs. Over the past decade, numerous randomized controlled trials (RCTs) have investigated possible pro-cognitive effects of various biological and psychological interventions [5–7]. Most pharmacological and neurostimulation interventions show no or only preliminary cognitive benefits, and cognitive remediation produce overall small-to-moderate effects on cognition [5–7]. Given this limited efficacy of monotherapies, multimodal interventions may be needed to produce a robust, enduring improvement in cognition [5]. Another major challenge is that limited or no transfer to everyday functioning is observed for the trials showing treatment-related cognitive benefits [5–7]. This highlights strategies to aid transfer as a key treatment priority.

Converging evidence from clinical, preclinical, and in-vitro studies suggests that cognitive impairments in MD originate from deficits in neuroplasticity, i.e., rewiring of neural connections in response to environmental challenges and input [8]. For example, substantial decreases in neuroprotective factors like brain-derived neurotrophic factor (BDNF) in the hippocampus and prefrontal cortex (PFC), which are essential brain areas for higher cognition, including memory and executive function, were found in rodent and post-mortem studies [8]. At a systemic level, magnetic resonance imaging (MRI) studies have documented hypo-activity in key nodes of the cognitive control network, including the dorsal PFC (dPFC), and volumetric changes in the hippocampus and dPFC in cognitively impaired relative to cognitively normal individuals with MD [9]. Novel, multimodal treatment strategies that directly target these underlying deficits in neuroplasticity and associated brain abnormalities are therefore a promising avenue to improve cognition in MD.

Controlled hypoxic exposure is a novel treatment strategy for increasing neuroplasticity and improving cognition. Human neurobiological adaptations to hypoxia are evolutionarily conserved and function as protective mechanisms that preserve central nervous system integrity under reduced oxygen availability, such as at high altitudes [10, 11]. At the core of this adaptive response is hypoxia-inducible factors (HIFs), which orchestrates cellular adaptation to hypoxia by upregulating the expression of key proteins implicated in cognitive function, including erythropoietin (EPO) and vascular endothelial growth factor (VEGF) [10–13]. The HIF-response forms a potential explanation of the ancient observation that residing in mountain sanctuaries has therapeutic effects in people with mental illness: Could low ambient oxygen at high altitudes mediate these benefits [14]? In a recent systematic review, we identified consistent neuroprotective effects and possible cognitive benefits of repeated exposure to 10-16% O_2_ over 30-240 min across humans and animals [15]. Some studies indicate that these effects are amplified when hypoxia is combined with motor-cognitive training, possibly through synergistic effects on neuroplasticity [12, 16]. However, conclusions were limited by small-scale trials and high risk of bias across previous human studies [15]. No randomized trials have investigated pro-cognitive effects of hypoxia in a psychiatric population, but a recent single-arm pilot study reported that hypoxia with concurrent motor-cognitive training was well-tolerated across mood and autism spectrum disorders, encouraging further studies [17].

The present outcome-assessor-blinded RCT investigated the effects of three weeks of daily 3.5-hour exposure to moderate normobaric hypoxia (12% O_2_) combined with cognitive training (H-CT) compared to treatment as usual (TAU) in cognitively impaired individuals with fully or partially remitted MD. This trial was based on encouraging findings from previous studies in mice [16] and conducted in parallel with a larger, double-blinded RCT in healthy individuals with four experimental arms (12% or 20% O_2_ (normoxia) with cognitive or sham training) that enabled mechanistic insights into the separate and combined effects of moderate hypoxia and cognitive training [18]. In this trial, we hypothesized that H-CT would produce robust and sustained improvements in broad cognition (primary outcome), particularly in executive functions, and normalization of working memory-related dPFC activity lasting at least one month after treatment completion. We also explored potential effects on everyday functioning, self-reported cognition, and quality of life. Using positron emission tomography (PET), we investigated the effects of H-CT on SV2A [^11^C]UCB-J binding as a readout for presynaptic density [19] in the hippocampus and frontal cortex. Finally, blood sampling was conducted to examine treatment effects on peripheral biomarkers of neuroplasticity, EPO, BDNF, and VEGF.

## Materials and Methods

### Study design

The trial had a randomized, controlled, outcome-assessor-blind, parallel group, superiority design. The study was conducted at the Department of Psychology, University of Copenhagen and Psychiatric Centre Copenhagen, Frederiksberg Hospital, and neuroimaging was conducted at the Copenhagen University Hospital, Rigshospitalet, Denmark. The study was approved by the Committee on Health Research Ethics (H-22028111) and the Danish Data Protection Agency (P-2022-354) in the Capital Region of Denmark, and pre-registered at ClinicalTrials.gov (NCT06121206) (open-access trial protocol: [20]). Given the highly mechanism- and hypothesis-driven nature of the H-CT intervention, patient and public involvement in study design and reporting was not judged feasible.

### Participants and recruitment

Participants were recruited through outpatient centers at the Mental Health Services of the Capital Region of Denmark, referrals from private practice psychiatrists, and online advertisements. Participants were somatically healthy 18-65-year-old individuals diagnosed with MD (bipolar disorder (BD) or major depressive disorder (MDD)) in full or partial remission, presenting with objective and/or subjective cognitive impairment. See **Supplementary Materials** for detailed in- and exclusion criteria, including criteria for healthy controls (HC) included for neuropsychological test score standardization and healthy individuals from our parallel trial included for routine blood value comparisons [18, 20]. After inclusion of the 60 participants (see power calculation below), we enrolled an additional four participants allocated to the H-CT group to obtain adequate statistical power for the PET analysis, as the scanner was temporarily unavailable early in the trial period (**Supplementary Materials**). All participants provided written informed consent.

### Randomization and masking

Randomizations to H-CT or TAU were stratified for age (< or ≥35 years) with a 1:1 allocation ratio. Blocked, random allocation sequence was created by an independent researcher through www.sealedenvelope.com. The sequence was inaccessible to the researchers conducting the randomizations through the REDCap online system. Participants were randomized upon inclusion and immediately informed about group allocation, which was necessary to allow for planning and timing of the time-extensive intervention (3.5 hours, 5-6 days per week for three weeks immediately following baseline assessment). Outcome assessors were blind to group allocations, and participants were instructed not to disclose their assigned intervention group during assessments.

### Procedure

Participants underwent screening for inclusion and exclusion criteria and were randomized to H-CT or TAU. We then conducted baseline assessments (week 0) of neurocognition (including estimation of IQ with the Danish Adult Reading Test) and functional MRI (fMRI) followed by a three-week intervention period. Participants were assessed in the week after the intervention (week 4) with a repeat of the neuropsychological test battery and, and a subsample also underwent PET with synaptic vesicle glycoprotein 2A (SV2A) radioligand [^11^C]UCB-J. After one month, neuropsychological assessments and fMRI were repeated (week 8). Routine blood samples were collected at inclusion, before treatment on day 8 (H-CT and healthy individual groups only) and day 19 of the intervention, and at week 8. Research blood samples to be analyzed for EPO, BDNF, and VEGF were collected before treatment at days 1 and 19. For ethical reasons, participants in the TAU group were offered the H-CT intervention after their week 8 follow-up visit. Treatment followed the same protocol as the intervention group, except that research blood samples were not taken at day 1 or 19. These participants were re-assessed with neuropsychological testing following treatment completion for exploratory within-group analyses.

### Intervention groups

#### Hypoxia with cognitive training

The H-CT group completed three weeks of 3.5-hour training sessions, 5-6 days per week (16 sessions total). The intervention took place in a sealed 20 m^2^ treatment room, into which fresh air with reduced O_2_-levels was blown by a 4-kW air compressor with a safety-approved system. Participants entered the room at 16% O_2_ (≈2,200m altitude), and oxygen levels were then reduced to 12% (≈4,400m altitude) over *M*±SD=47±5 minutes. Participants wore a pulse oximeter which measured pulse and blood oxygen saturation (SpO_2_) and completed the Environmental Symptoms Questionnaire (ESQ) after each treatment session to systematically investigate symptoms of altitude sickness. Adverse events (AEs) were recorded, and serious AEs were reported to the overseeing ethics committee. During treatment sessions, participants completed cognitive training on the HappyNeuron Pro online platform interleaved with small breaks inside the altitude room. See **Supplementary Materials** and trial protocol for details on the treatment sessions [20].

#### Treatment as usual

Treatment as usual entailed an eight-week period where participants completed assessments at the same intervals as the H-CT group (except routine blood sampling on day 8). They were not systematically in contact with study personnel except for during assessment visits.

### Primary outcome

The pre-registered primary outcome was change in the ‘speed of complex cognitive processing’ cognitive composite score from baseline to treatment completion, as this composite score previously showed improvement after high-dose EPO vs. saline infusions [20, 21]. The composite score was created by averaging *z*-transformed cognitive measures of sustained attention, memory, and executive functions, all commonly used to assess cognition in MD [22]: Rey Auditory Verbal Learning Test (RAVLT) lists I-V total recall, Repeatable Battery for the Assessment of Neuropsychological Status (RBANS) Coding, Verbal Fluency with the letter “D”, Wechsler Adult Intelligence Scale (WAIS)-III Letter-Number Sequencing (LNS), Trail Making Test Part B (TMT-B), and Rapid Visual Information Processing (RVP) mean reaction time (Cambridge Neuropsychological Test Automated Battery; CANTAB, Cambridge Cognition Ltd.). The composite score is on a *z*-scale, so any treatment effects can be interpreted as differential change in SD units between the groups.

### Secondary, tertiary, and mechanistic outcomes

The secondary cognition outcome was change in the ‘mean choices to correct’ score in the One-Touch Stockings of Cambridge (OTS) task from CANTAB (executive functions) from baseline to treatment completion. This was based on our previous trial which showed improvements in this measure following HappyNeuron Pro training [23]. The secondary functioning outcome was change in cognitive skills in a real-life environment (i.e., cooking a meal in a kitchen) measured with the Cognitive Assessment in Virtual Reality (CAVIR) ‘global composite’ measure from baseline to one-month follow-up (not assessed at treatment completion), based on International Society for Bipolar Disorders (ISBD) recommendations to include a measure of functioning as a secondary trial outcome [22]. Finally, the secondary neuroimaging outcome was change in dPFC activity during working memory measured with an fMRI N-back task from baseline to one-month follow-up. The specific measure was mean percent blood-oxygen-level-dependent (BOLD) signal change during general working memory from a right dorsolateral prefrontal cortex (dlPFC) region of interest (ROI) which has shown response to EPO treatment [24].

Tertiary outcomes included global cognition and domain-specific cognitive function based on the full neuropsychological test battery as well as clinician-rated and self-reported measures of functioning, subjective cognition, quality of life, and sleep quality.

Mechanistic outcomes included PET [^11^C]UCB-J non-displaceable binding potential (*BP_ND_*) in the hippocampus and frontal cortex at treatment completion. Change in working memory-related BOLD response across the whole dPFC from baseline to follow-up were also assessed. Finally, treatment effects on peripheral neuroplasticity biomarkers, EPO, BDNF, and VEGF, from prior to day 1 and day 19 of treatment were assessed. For a full list of the tertiary and mechanistic outcomes and domain calculations, see **Supplementary Materials**.

### PET analysis

Details on PET acquisition, preprocessing, and quantification are available in the **Supplementary Materials**. Following a six min transmission scan, participants underwent a 90 min emission scan that began at the time of intravenous [^11^C]UCB-J bolus injection over ∼20 seconds. Images were motion corrected using the Automated Image Registration (AIR) software with the reconcile command (v. 5.2.5) [25]. PET images were co-registered to each participant’s T1-weighted MR image acquired at follow-up, and tissue time-activity curves were extracted from automatically defined ROIs [26–28] using the PVElab software pipeline (https://nru.dk/pveout/). Quantification was performed using a simplified reference tissue model 2 (SRTM2) to estimate *BP_ND_*, using the centrum semiovale (white matter) as reference region [29]. Our a priori ROIs were the hippocampus and frontal cortex, but we also extracted estimated *BP_ND_* for other cortical and subcortical regions to explore possible group differences across the whole brain (**Supplementary Materials**).

### fMRI analysis

The fMRI paradigm was a spatial working memory N-back task. Details on the paradigm, fMRI data acquisition, preprocessing, subject-level analysis, and in-scanner behavioral data are available in the **Supplementary Materials**. In brief, the task was modelled at the subject-level with three contrasts: 2-back>0-back, 2-back>1-back, and 1-back>0-back to model general working memory, high-load specific, and low-load specific working memory, respectively. We constructed an eight mm spherical right dlPFC ROI around x=40, y=34, z=29 and extracted the mean percent BOLD signal change for statistical analyses. To explore neural effects of H-CT in the broader dPFC area and whole brain, we also conducted two-way mixed effects analyses of variance in FMRI Expert Analysis Tool (FEAT). Here, we used the 2-back>0-back, 2-back>1-back, and 1>0-back contrasts and used FMRIB’s Local Analysis of Mixed Effects (FLAME) for estimations. We set the significance level to *p*<0.05 and corrected for multiple comparisons with a cluster-forming threshold of *Z*=2.56 (*p*<0.005). For the dPFC analysis, we defined a volume of interest (VOI) which included the bilateral superior and medial frontal gyri, the frontal poles, and the superior parts of the anterior division of the cingulate gyrus on the standard Montreal Neurological Institute (MNI) template in FMRIB Software Library (FSL). For clusters showing treatment-related changes in activity over time, we extracted mean percent BOLD signal change for correlation analyses with cognition.

### Biochemistry analysis

Routine blood samples were analyzed immediately for hemoglobin, erythrocyte, reticulocyte, and thrombocyte counts at the Department of Clinical Biochemistry, Copenhagen University Hospital. Research blood samples were stored at -80 ᵒC at Frederiksberg Hospital until analysis. Serum concentrations of EPO (U-PLEX, #K151VXK-2), VEGF (V-PLEX, #K151RHD-2), and BDNF (U-PLEX, #K151WK-2) were quantified using the Meso Scale Discovery (MSD) electrochemiluminescence immunoassay platform. Measurements were performed on a SECTOR Imager 6000 plate reader according to the manufacturer’s instructions (Meso Scale Diagnostics, Rockville, MD, USA). Data acquisition and analysis were conducted using MSD Workbench software. Analyses were performed in duplicate, and results are reported as mean values. The laboratory technician was blinded to sample identity.

### Statistical analysis

A power calculation based on data from our previous RCT of EPO infusions [20, 21] showed that 52 participants (26 per group) would achieve 80% power to detect a greater improvement of 0.40 or more *z*-scores (moderate effect size) in H-CT vs. TAU for the primary outcome (‘speed of complex cognitive processing’ composite *z*-score) at an α-level of 0.05 [20]. To accommodate for dropouts, we aimed to recruit 30 participants per group.

All outcome analyses were performed in R (v.2026.01.0). See **Supplementary Materials** for details on score standardization and domain calculations. All participants with baseline data were included in the analyses of primary, secondary, and tertiary outcomes. For these, we performed baseline-constrained linear mixed models (LMMs) with an unstructured covariance pattern. Fixed effects were stratum (age:< or ≥35 years), time, treatment, and their interaction (time*treatment). The significance level was set to *p*<0.05 (two-tailed). We applied Benjamini-Hochberg (BH) corrections to control for 5% false discovery rate for secondary and tertiary outcomes. Significant results were followed up by post hoc mood- and diagnosis-corrected analyses. Similar LMMs were performed for the mechanistic blood-based biomarkers (where time: day 1 and day 19). Group differences in estimated [^11^C]UCB-J *BP_ND_* were assessed with analyses of covariance (ANCOVAs), including stratum as covariate. We did not correct for multiple comparisons due to the exploratory purpose of these group comparisons in *BP_ND_*.

## Results

### Participant flow and missing data

See **Figure 1** for the CONSORT flow diagram. Between April 12, 2023, and June 4, 2025, 70 individuals were screened; 64 participants were enrolled (28 BD; 36 MDD) and randomized to H-CT (*n*=34) or TAU (*n*=30). Final follow-up was completed on July 30, 2025. Three participants (before baseline: one H-CT, one TAU; after baseline: one H-CT) withdrew prior to treatment. The remaining 29 TAU participants completed the follow-up assessments. In the H-CT group, six discontinued during treatment (see **Figure 1** for reasons); 26 completed follow-up assessments. Dropout rates were 24% (H-CT), 3% (TAU), and 14% overall. The analyses included all randomized participants with baseline data (33 H-CT, 29 TAU). For the PET imaging analysis, data from 28 participants (13 H-CT, 15 TAU) were analyzed (flow diagram in **Figure S1**). See **Supplementary Materials** for missing fMRI and PET data. Eighteen TAU participants completed H-CT and exploratory follow-up after their initial one-month follow-up.

**Figure 1.**
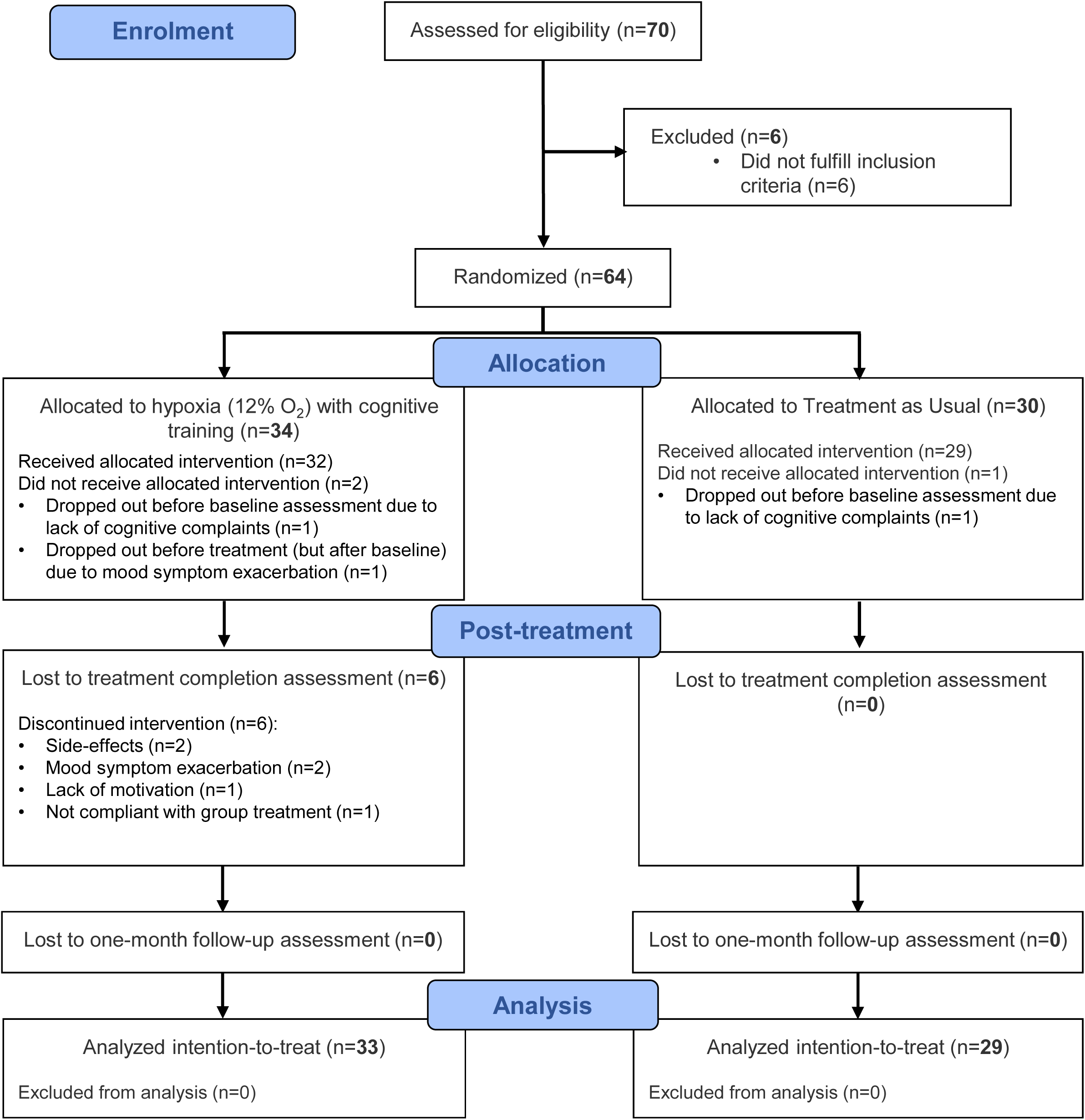
CONSORT Flow diagram.

### Sample characteristics

See **Table S1** for baseline demographic and clinical characteristics of the full sample. The sample had a median age of 35 years (IQR=29-53) and were predominantly female (69%). Two-thirds were employed or studying, and subsyndromal mood symptom levels were low. The sample was evenly distributed between BD (44%) and MDD (56%), and 77% received psychotropic medication. The treatment groups were comparable across demographic and clinical variables (**Table 1**). Participants with MD were matched to HC on age, sex, and IQ (*p*s≥0.12) but scored lower on all cognitive measures (*p*s≤0.06) except verbal memory (*p*=0.38), and facial emotion recognition (*p*=0.32) (**Table S1**).

**Table 1.**
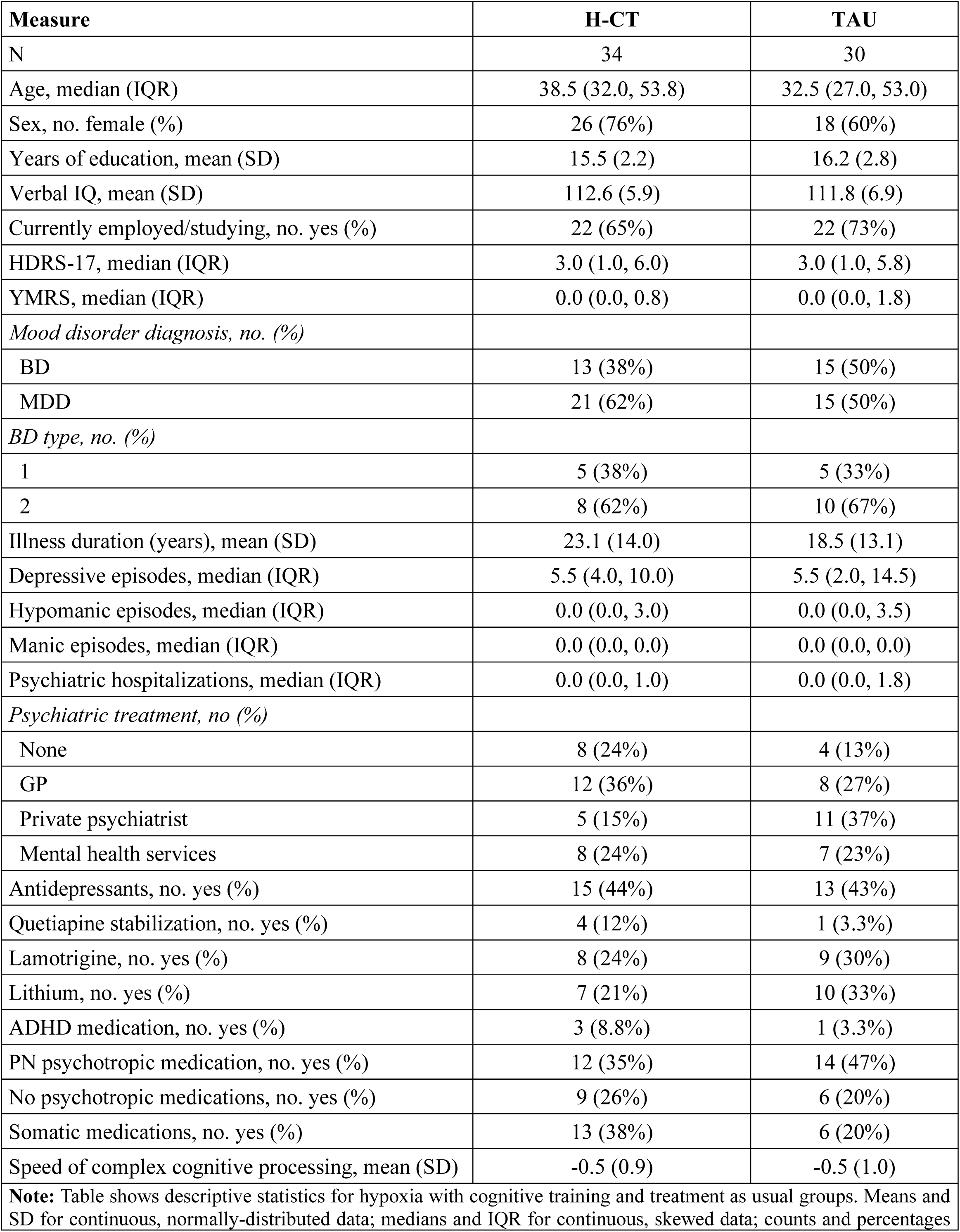
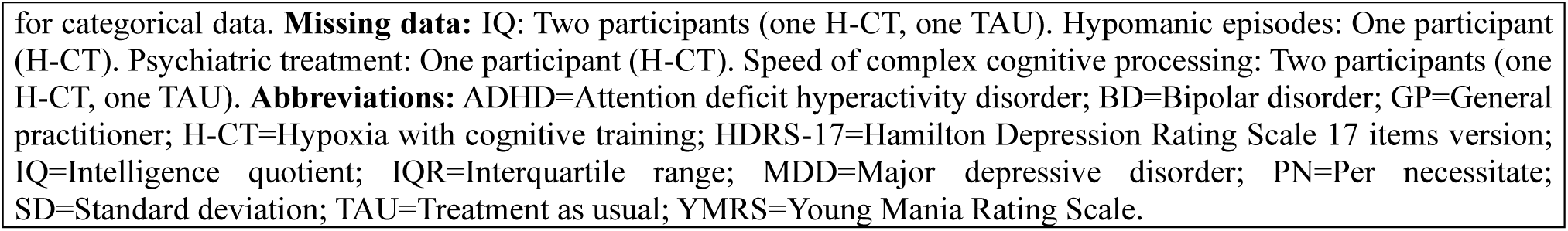
Demographic and clinical characteristics in hypoxia with cognitive training and treatment as usual groups.

| Measure | H-CT | TAU |
| --- | --- | --- |
| N | 34 | 30 |
| Age, median (IQR) | 38.5 (32.0, 53.8) | 32.5 (27.0, 53.0) |
| Sex, no. female (%) | 26 (76%) | 18 (60%) |
| Years of education, mean (SD) | 15.5 (2.2) | 16.2 (2.8) |
| Verbal IQ, mean (SD) | 112.6 (5.9) | 111.8 (6.9) |
| Currently employed/studying, no. yes (%) | 22 (65%) | 22 (73%) |
| HDRS-17, median (IQR) | 3.0 (1.0, 6.0) | 3.0 (1.0, 5.8) |
| YMRS, median (IQR) | 0.0 (0.0, 0.8) | 0.0 (0.0, 1.8) |
| *Mood disorder diagnosis, no. (%)* |  |  |
| BD | 13 (38%) | 15 (50%) |
| MDD | 21 (62%) | 15 (50%) |
| *BD type, no. (%)* |  |  |
| 1 | 5 (38%) | 5 (33%) |
| 2 | 8 (62%) | 10 (67%) |
| Illness duration (years), mean (SD) | 23.1 (14.0) | 18.5 (13.1) |
| Depressive episodes, median (IQR) | 5.5 (4.0, 10.0) | 5.5 (2.0, 14.5) |
| Hypomanic episodes, median (IQR) | 0.0 (0.0, 3.0) | 0.0 (0.0, 3.5) |
| Manic episodes, median (IQR) | 0.0 (0.0, 0.0) | 0.0 (0.0, 0.0) |
| Psychiatric hospitalizations, median (IQR) | 0.0 (0.0, 1.0) | 0.0 (0.0, 1.8) |
| *Psychiatric treatment, no (%)* |  |  |
| None | 8 (24%) | 4 (13%) |
| GP | 12 (36%) | 8 (27%) |
| Private psychiatrist | 5 (15%) | 11 (37%) |
| Mental health services | 8 (24%) | 7 (23%) |
| Antidepressants, no. yes (%) | 15 (44%) | 13 (43%) |
| Quetiapine stabilization, no. yes (%) | 4 (12%) | 1 (3.3%) |
| Lamotrigine, no. yes (%) | 8 (24%) | 9 (30%) |
| Lithium, no. yes (%) | 7 (21%) | 10 (33%) |
| ADHD medication, no. yes (%) | 3 (8.8%) | 1 (3.3%) |
| PN psychotropic medication, no. yes (%) | 12 (35%) | 14 (47%) |
| No psychotropic medications, no. yes (%) | 9 (26%) | 6 (20%) |
| Somatic medications, no. yes (%) | 13 (38%) | 6 (20%) |
| Speed of complex cognitive processing, mean (SD) | -0.5 (0.9) | -0.5 (1.0) |
| **Note:** Table shows descriptive statistics for hypoxia with cognitive training and treatment as usual groups. Means and SD for continuous, normally-distributed data; medians and IQR for continuous, skewed data; counts and percentages |  |  |

for categorical data. **Missing data:** IQ: Two participants (one H-CT, one TAU). Hypomanic episodes: One participant (H-CT). Psychiatric treatment: One participant (H-CT). Speed of complex cognitive processing: Two participants (one H-CT, one TAU). **Abbreviations:** ADHD=Attention deficit hyperactivity disorder; BD=Bipolar disorder; GP=General practitioner; H-CT=Hypoxia with cognitive training; HDRS-17=Hamilton Depression Rating Scale 17 items version; IQ=Intelligence quotient; IQR=Interquartile range; MDD=Major depressive disorder; PN=Per necessitate; SD=Standard deviation; TAU=Treatment as usual; YMRS=Young Mania Rating Scale.

### Treatment adherence and safety

The *n*=26 participants from the H-CT group who completed the intervention attended a median of 15 (IQR=15-15.8) of 16 treatment sessions and spent *M*±SD=27±4 hours in total on cognitive training. The intervention led to the expected drop in SpO_2_ with a median of 85% (IQR=83%-87%), compared to 97.5% (IQR=97%-98%) in the healthy participant normoxia group from our parallel study [18].

Two H-CT participants experienced mood symptom exacerbation (one depressive, one manic) during the intervention that led to dropout from the study, which was classified as serious AEs unrelated to the intervention. Four other (non-severe) AEs in H-CT included dizziness (*n*=2, one H-CT; one TAU participant during H-CT), nausea (one TAU participant during H-CT), and headache (one H-CT), which all led to treatment cessation. There were no AEs during TAU. The H-CT participants who completed the intervention reported low levels of side-effects on the ESQ (median daily score=4, IQR=1-5; score range: 0-55). There was no significant change in hemoglobin, erythrocytes, reticulocytes, or thrombocytes during the intervention in the H-CT group compared to healthy individuals undergoing normoxia or TAU (*p*s≥0.12). Details on treatment adherence, tolerability, and routine blood values for each group and timepoint are provided in **Supplementary Materials** (incl. **Tables S2-S4** and **Figure S2**).

### Primary outcome

**Table 2** shows results from the primary and secondary outcomes. There was no statistically significant effect of H-CT compared to TAU on ‘speed of complex cognitive processing’ at treatment completion or one-month follow-up (*p*s≥0.24) (**Figure 2A**).

**Figure 2.**
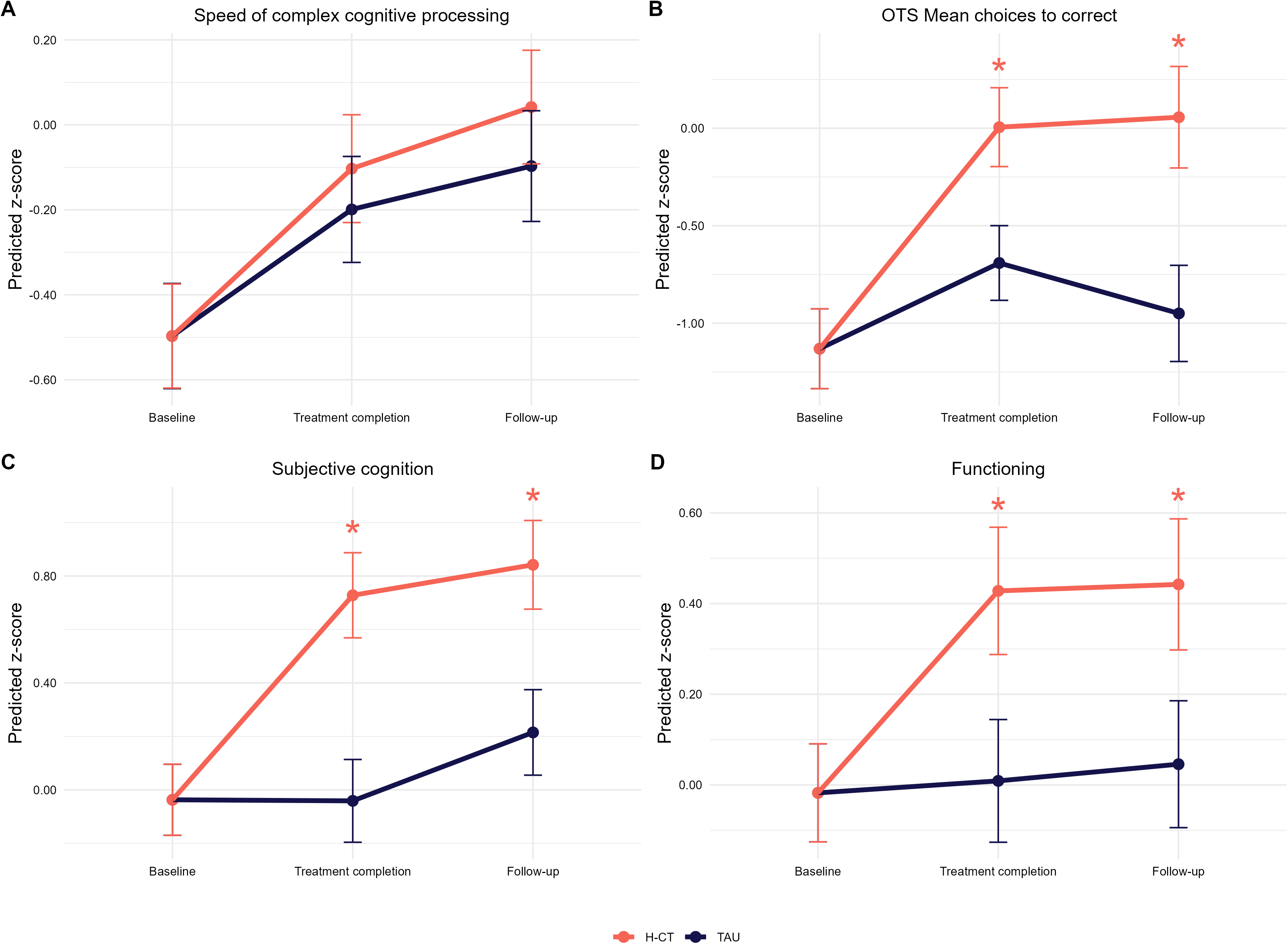
Treatment effects in primary outcome (A), secondary cognitive outcome (B), subjective cognition (tertiary outcome, C), and functioning (tertiary outcome, D). Stars mark significant improvements in H-CT group compared to TAU group (uncorrected). Speed of complex cognitive processing composite score consisted of averaged *z*-transformed (and inversed when relevant) values from Rey Auditory Verbal Learning Test (RAVLT) lists I-V total recall, Repeatable Battery for the Assessment of Neuropsychological Status (RBANS) Coding, Verbal Fluency with the letter “D”, Wechsler Adult Intelligence Scale (WAIS)-III Letter-Number Sequencing (LNS), Trail Making Test Part B (TMT-B), and Rapid Visual Information Processing (RVP; CANTAB) mean reaction time. Subjective cognition score was *z*-transformed and inversed Cognitive Complaints in Bipolar Disorder Rating Assessment (COBRA) scores. Functioning composite score consisted of averaged *z*-transformed and inversed values from Functional Assessment Short Test (FAST) (baseline and one-month follow-up only), Work and Social Adjustment Scale (WSAS), and Sheehan Disability Scale (SDS). **Abbreviations:** H-CT=Hypoxia with cognitive training; OTS=One-touch stockings of Cambridge (CANTAB); TAU=Treatment as usual.

**Table 2.** Primary and secondary outcomes.

|  |  | Treatment completion |  |  | One-month follow-up |  |  |
| --- | --- | --- | --- | --- | --- | --- | --- |
|  | Group | Treatment effect | 95% CI | p-value (adj.) | Treatment effect | 95% CI | p-value (adj.) |
| **Primary outcome** |  |  |  |  |  |  |  |
| Speed of complex cognitive processing | H-CT | 0.10 | -0.10; 0.30 | 0.30 | 0.15 | -0.10; 0.40 | 0.24 |
|  | TAU | - | - | - | - | - | - |
| **Secondary cognitive outcome** |  |  |  |  |  |  |  |
| OTS Mean choices to correct | H-CT | 0.76 | 0.23; 1.28 | **0.01 (0.02)** | 1.07 | 0.36; 1.77 | **0.004 (0.02)** |
|  | TAU | - | - | - | - | - | - |
| **Secondary neuroimaging outcome** |  |  |  |  |  |  |  |
| Mean percent BOLD signal change in dorsal prefrontal activity during working memory | H-CT | - | - | - | 0.09 | -0.25; 0.43 | 0.61 |
|  | TAU | - | - | - | - | - | - |
| **Secondary functioning outcome** |  |  |  |  |  |  |  |
| CAVIR global composite | H-CT | - | - | - | -0.12 | -0.51; 0.27 | 0.54 |
|  | TAU | - | - | - | - | - | - |
| **Note:** Table shows results from linear mixed models comparing H-CT and TAU from baseline to treatment completion and one-month follow-up. Speed of complex cognitive processing composite score consisted of averaged z-transformed (and inversed when relevant) values from RAVLT lists I-V total recall, RBANS Coding, Verbal Fluency with the letter “D”, WAIS-III LNS, TMT-B, and RVP mean reaction time. CAVIR global composite consisted of averaged z-transformed values from the five subtests of the task. **Sample sizes:** Baseline: 33 H-CT, 29 TAU. Treatment completion: 26 H-CT, 29 TAU. One-month follow-up: 26 H-CT, 29 TAU. **Missing data:** Secondary neuroimaging outcome (see Supplementary Materials for details on missing functional magnetic resonance imaging data): 10 participants at baseline (seven H-CT, three TAU), eight participants at follow-up (five H-CT, three TAU). CAVIR global composite: one participant (TAU) at follow-up. **Abbreviations:** Adj.=Adjusted; BOLD=Blood-oxygen-level-dependent; CAVIR=Cognitive Assessment in Virtual Reality; CI=Confidence interval; H-CT=Hypoxia with cognitive training; OTS=One-touch Stockings of Cambridge; RAVLT=Rey Auditory Verbal Learning Test; RBANS=Repeatable Battery for the Assessment of Neuropsychological Status; RVP=Rapid Visual Information Processing (CANTAB); TAU=Treatment as usual; TMT=Trail Making Test; WAIS-III LNS=Wechsler Adult Intelligence Scale III Letter-Number Sequencing. |  |  |  |  |  |  |  |

### Secondary outcomes

There was a statistically significant beneficial effect of H-CT compared to TAU on the secondary cognitive outcome, OTS mean choices to correct, with a large effect size at treatment completion, which prevailed at the one-month follow-up (treatment completion: treatment effect=0.76, adj. *p*=0.02; one-month follow-up: treatment effect=1.07, adj. *p*=0.02) (**Figure 2B**). This effect survived adjustment for mood symptoms and diagnosis at both timepoints (treatment completion: treatment effect=0.85, *p*=0.002; one-month follow-up: treatment effect=1.15, *p*=0.003). There were no effects on working memory-related dlPFC activity (*p*=0.61) or the CAVIR assessment of functioning (*p*=0.54) at one-month follow-up.

### Tertiary outcomes

**Table 3** shows results from the tertiary outcomes. The H-CT group displayed reduction in self-reported cognitive difficulties (treatment completion: treatment effect=0.77, adj. *p*=0.002; one-month follow-up: treatment effect=0.62, adj. *p*=0.02) and reported improved daily functioning (treatment completion: treatment effect=0.41, adj. *p*=0.049; one-month follow-up: treatment effect=0.40, adj. *p*=0.049) (**Figure 2C+2D**). The H-CT group also showed improved executive functions at domain-level compared to TAU at one-month follow-up, which rendered non-significant after adjustment (treatment effect=0.33, adj. *p*=0.12). At treatment completion, the H-CT group displayed improvements on quality of life (treatment effect=0.46, adj. *p*=0.03) and improved facial emotion recognition accuracy that rendered non-significant after adjustment (treatment effect=0.35, adj. *p*=0.12) compared to TAU. All significant results prevailed after adjusting for mood symptoms and diagnosis (*p*s≤0.05). There were no significant correlations between changes in the secondary outcome/executive functions and changes in subjective cognition/functioning/mood (**Table S5**) or treatment dose (**Table S6**). See **Supplementary Materials** for LMM results on individual measures (**Table S7**) and analyses in TAU participants who completed H-CT (**Table S8**).

**Table 3.**
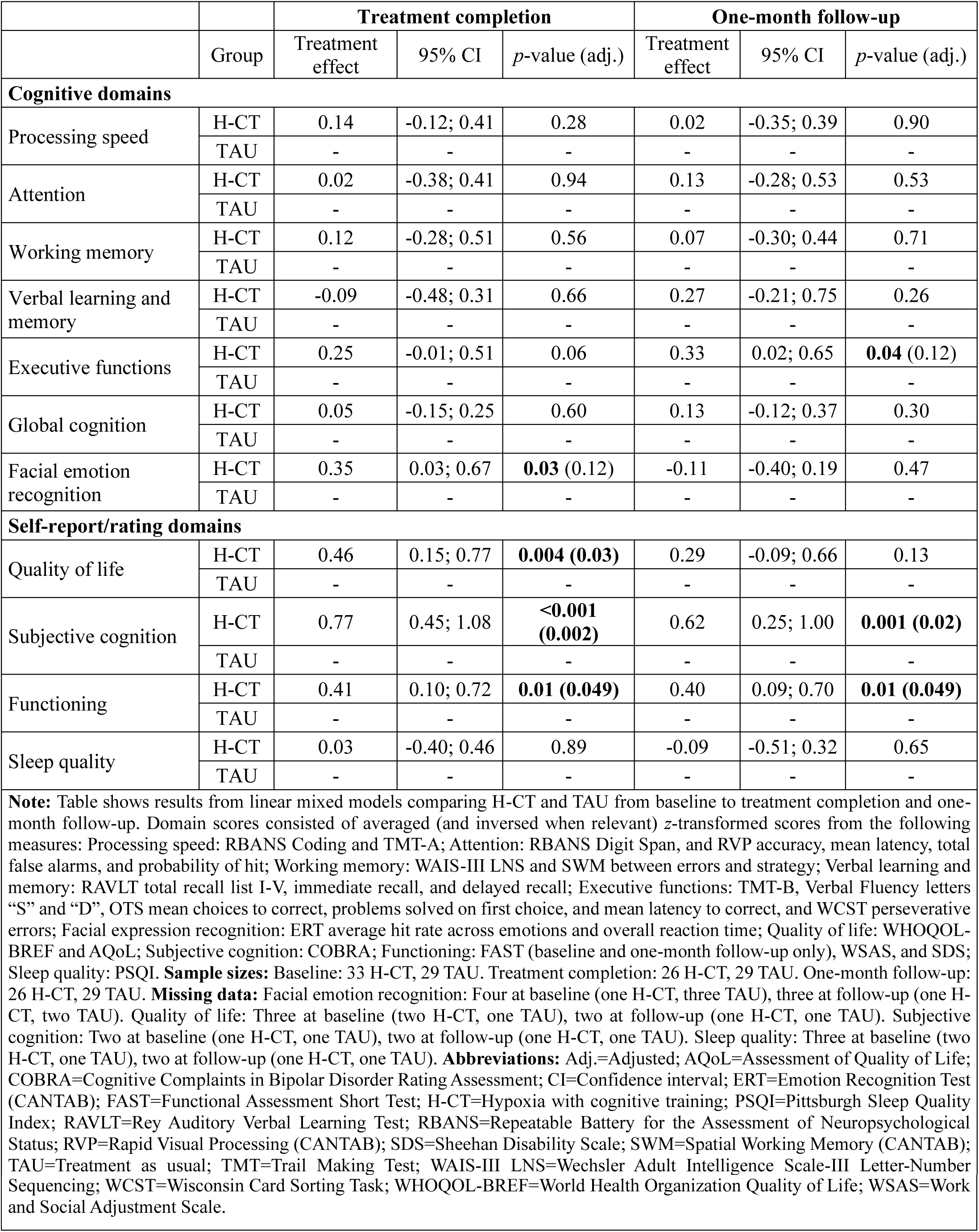
Tertiary outcome domains.

### Mechanistic analyses

#### Presynaptic density

There were no group differences in [^11^C]UCB-J *BP_ND_* in the hippocampus or frontal cortex for H-CT vs. TAU at treatment completion (*ps*≥0.16). However, exploratory analyses showed a significant lower *BP_ND_* in the nucleus accumbens for H-CT vs. TAU (H-CT: *M*±SD=4.56±0.48, TAU: *M*±SD=5.00±0.66, unadj. *p*=0.04, effect size(partial eta-squared)=0.14) (**Figure 3A**). A post hoc Pearson correlation analysis showed a significant correlation between lower *BP_ND_* in the nucleus accumbens and greater improvements in OTS mean choices to correct (secondary outcome) from baseline to treatment completion (*r*=-0.42, *p*=0.03) (**Figure 3B**), but no correlation with change in mood symptoms or any of the other cognitive or functioning outcomes showing H-CT-related improvements (*ps*≥0.07). There were no group differences in the remaining exploratory ROIs (**Table S9**).

**Figure 3.**
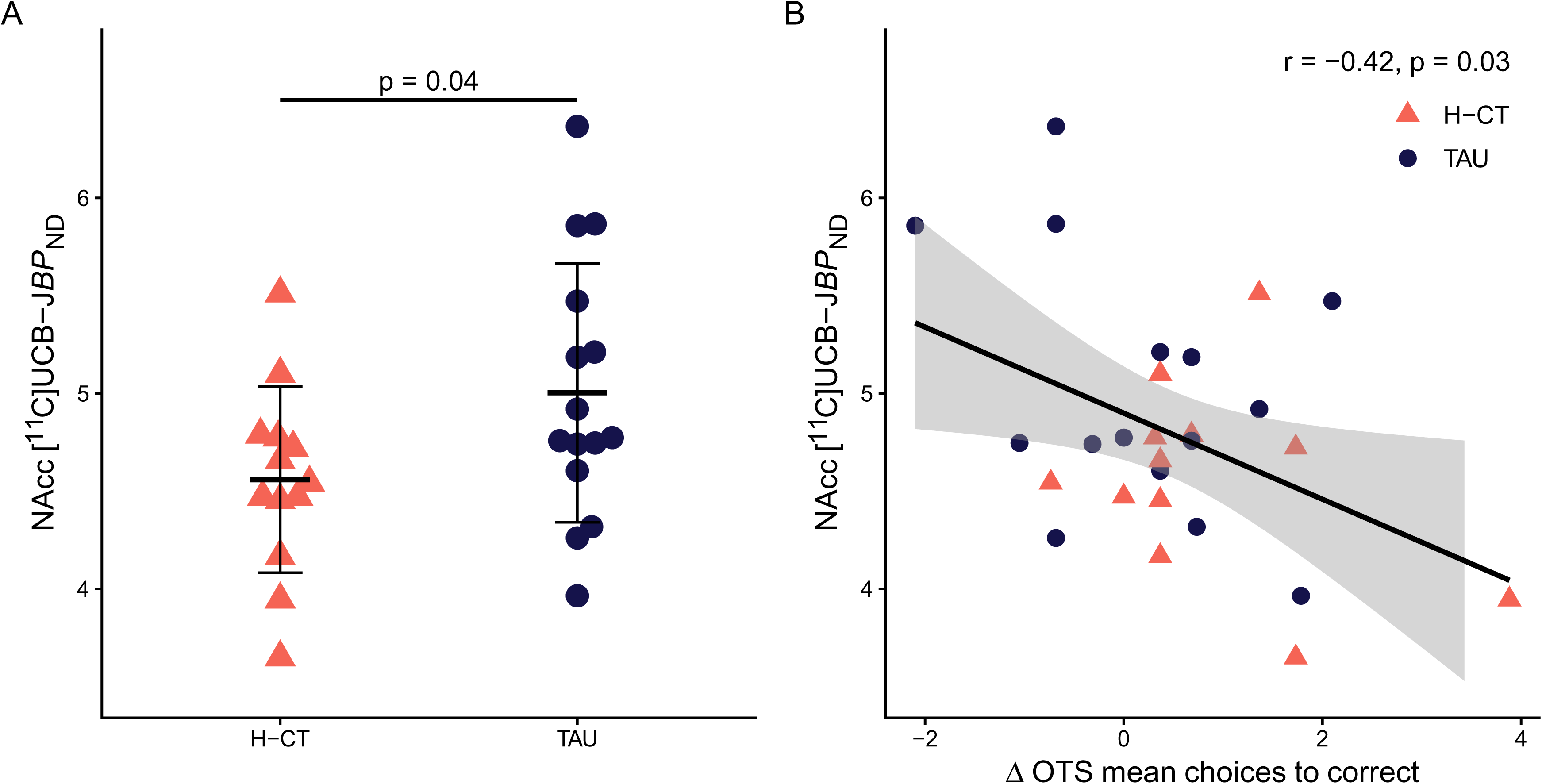
Exploratory treatment effects on presynaptic density in the nucleus accumbens at treatment completion. A: Comparison of synaptic vesicle glycoprotein 2A (SV2A) [^11^C]UCB-J binding in the nucleus accumbens after three weeks of H-CT (*n*=13) versus TAU (*n*=15). [^11^C]UCB-J non-displaceable binding potential (*BP_ND_*) was quantified using a simplified reference region model 2 (SRTM2) with centrum semiovale (white matter) as reference. The crossbars show the means and standard deviations for each group. B: Association between nucleus accumbens *BP_ND_* and improvements in OTS mean choices to correct (secondary outcome) from baseline to treatment completion across both groups (*n*=28). The shaded grey area represents the 95% confidence intervals. There was no significant correlation with improvement in this measure from baseline to one-month follow-up (data not shown). **Abbreviations**: H-CT=Hypoxia with cognitive training; NAcc=Nucleus accumbens; OTS=One-touch stockings of Cambridge (CANTAB); TAU=Treatment as usual.

#### Working memory-related neural activity

Within the dPFC VOI, time-by-treatment interaction analyses revealed greater deactivation from baseline to follow-up in H-CT vs. TAU in the right precentral gyrus (*p*=0.002) and left precentral gyrus (*p*=0.03) during the 1>0-back contrast (low-load working memory) **(Figure S3**). Post hoc analysis revealed that improvements in working memory performance from baseline to follow-up correlated with more deactivation over time in the right precentral gyrus cluster (*p*=0.03). Consistent with this, three clusters at the whole-brain level demonstrated greater deactivation over time in H-CT vs. TAU during low-load working memory: right precentral gyrus (*p*<0.001), left postcentral gyrus (*p*<0.001), and right lateral occipital cortex (*p*<0.001). There was also a cluster in the right lateral occipital cortex with greater deactivation over time for H-CT vs. TAU in the 2>0-back contrast (general working memory; *p*=0.005) (**Table S10**). There were no significant correlations between H-CT-related improvement in executive function and change in neural activity in the clusters modulated by the intervention (*p*s≥0.14) (**Table S11**).

#### Peripheral biomarkers

There was no change in serum EPO, BDNF, or VEGF from day 1 to day 19 of the intervention for H-CT compared to TAU (*p*s≥0.10) (**Table S12**).

#### Mood symptoms

There was a reduction in subsyndromal mood symptoms in H-CT vs. TAU from before to immediately after the intervention, which rendered non-significant at follow-up (treatment completion: treatment effect=-3.22, 95% CI=[-5.53;-0.91], *p*=0.007; one-month follow-up: treatment effect=-1.39, 95% CI=[-4.86;2.07], *p*=0.42).

## Discussion

This outcome assessor-blinded, multimodal RCT in 64 cognitively impaired participants with MDD or BD in full or partial remission showed that daily moderate hypoxia combined with cognitive training over three weeks yields no significant improvement in the broad primary cognitive outcome. However, it produced a robust and sustained improvement in the secondary executive function outcome and tertiary measures of self-reported cognition, mood, and functioning. Mechanistic exploratory analyses revealed a lower presynaptic [^11^C]UCB-J binding in the nucleus accumbens in the week after H-CT versus TAU and greater dPFC and occipital deactivation during working memory one month after the intervention, respectively. There was a moderate (expected) dropout (24%) in the H-CT group, but the intervention was well-tolerated with minimal side-effects.

The long-lasting and robust H-CT-associated improvement in executive functions, measured with the OTS, suggests that daily moderate hypoxia combined with cognitive training over three weeks may produce lasting neuroplastic changes in MD, in line with the emerging evidence on benefits of moderate hypoxia [15] and demonstration of broad cognitive gains in our parallel, double-blind RCT in healthy volunteers [18]. Indeed, we observed that the executive function gains were even broader one month after H-CT completion, extending to several tests probing mental flexibility, attention switching, and verbal fluency. In addition to these cognitive improvements, the H-CT group showed lower presynaptic density in the week after H-CT completion within the nucleus accumbens- a region implicated in goal- and reward-oriented behavior [30–32] - an effect that correlated with greater improvement in executive functions (secondary trial outcome). This finding partly aligns with our parallel study in healthy individuals, where H-CT-related lower hippocampal *BP_ND_* was associated with greater cognitive gains and interpreted as enhanced adaptive synaptic pruning [33]. Interestingly, this direction of the association with *lower BP_ND_* being related to *greater* cognitive benefits differs from one previous [^11^C]UCB-J PET study in MDD that showed a link between lower presynaptic density in the anterior cingulate cortex and dlPFC and poorer verbal memory or working memory [34]. Moreover, the H-CT group showed decreased general and low-load working memory activity in the occipital cortex and dPFC compared with TAU at one-month follow-up, which may reflect long-lasting increased neural efficiency during cognitively challenging tasks [9]. Together, the executive function improvements observed immediately following H-CT accompanied by the lower presynaptic density, and the further executive function increase at one-month follow-up accompanied by working memory-related dPFC and occipital changes, may suggest a two-phase trajectory of neuroplastic adaptation. Early gains may reflect rapid synaptic responsiveness to hypoxia and cognitive training, whereas the accentuated effects at follow-up may indicate ongoing consolidation and the gradual emergence of neuroplastic changes at the systems level in the brain. This interpretation aligns with the concept of ‘hormesis’ which states that continued or repeated exposure to a mild stressor (i.e., hypoxia or intense cognitive training) over an extended period of time combined with adequate physiological recovery intervals can enhance bodily and cerebral functions [35]. Finally, the decrease in subsyndromal mood symptoms immediately following treatment is in line with our study of EPO injections in MDD [36] and could also signal increased neuroplasticity after H-CT [8].

In contrast to the treatment benefits on executive functions, we observed no effects of H-CT on the broad cognition outcome or on dlPFC activity during general working memory. In our parallel trial involving healthy individuals, H-CT improved broad cognition at the one-month follow-up alongside hypoxia-driven alterations in erythropoiesis and changes in serum levels of EPO and VEGF [33]. Conversely, in the current study, we observed no comparable changes in red blood cell indices or circulating EPO and VEGF despite the observed median SpO_2_ of 85% (IQR=83%-87%) in the H-CT group, indicating that hypoxia was delivered as intended. This unexpected divergence may reflect a fundamental deficit in the body’s physiological adaptation to hypoxia in MD, which may partially explain the attenuated response to H-CT and absence of broader cognitive benefits. In line with this interpretation, clinical studies have reported dysregulated peripheral and central VEGF signaling and compromised neurovascular responses in MDD - findings consistent with an inability to mount proper angiogenic or erythropoietic adaptations [13]. Collectively, this could suggest that deficient hypoxia-related adaptations, which underlie a range of somatic diseases [10], may also generally contribute to the neuroplastic maladaptation and cognitive impairments observed in MD [8]. Further, evidence from previous cognitive remediation trials shows that individuals living cognitively stimulating lives, e.g., through employment or studying, benefit more from interventions [37]. Indeed, employment rates were lower in MD than in our healthy participant trial (69% vs. 88%), which could also contribute to the more selective effects observed here [33].

The improvements in combined self- and clinician-rated functioning after H-CT makes the present study one of the few to demonstrate improved daily functioning after a pro-cognitive intervention in MD [5–7]. The functional enhancements could be linked to the accompanying improvements across executive functions, as this cognitive domain is strongly linked to functional abilities [3]. Our trial suggests that multimodal interventions with more intense brain stimulation (i.e., through hypoxia) than what is seen in unimodal interventions are necessary to produce cognitive effects that are large enough to translate to improved functioning. However, we found no direct correlation between improvements in executive functions and in daily-life functioning, which challenges the interpretation that these were mechanistically linked. Given that the functioning domain included self-reported scales, an expectancy bias could also have affected the result as participants were not blinded. Another possibility for the observed functional improvements could be that the intense regimen of the intervention provided structure and regulated social rhythms in the trial participants [38]. Our experience was that completing the program boosted self-efficacy and was highly rewarding for the participants. Together, these insights can be leveraged in future trial designs: intensive treatment regimens are likely necessary to ensure neuroplasticity upregulation and functional benefits, and this may be feasible if the treatment is relatively short-term (e.g., 3-4 weeks). However, treatments lasting 5-6 days per week are likely not ready for implementation due to high costs and interference with obligations in everyday life. Future trials should therefore investigate the minimal dose of H-CT necessary to produce cognitive effects and explore more mobile delivery options, such as oxygen masks [14].

To facilitate recruitment of study participants, prescreening for cognitive impairment involved either objective and/or subjective difficulties. When formally testing, 33% were not objectively cognitively impaired and that could explain the lack of effect on the primary outcome because objective cognitive impairment at baseline is the most consistent predictor of cognitive improvement in intervention trials [39]. Nevertheless, subjective cognitive impairment may reflect decline from premorbid functioning [40], making interventions clinically relevant. Further, our parallel healthy participants study showed cognitive improvements in unimpaired individuals [33]. Generalizability may be limited due to strict somatic exclusion criteria, which were necessary due to the novelty of the H-CT intervention. Further, our participants were not blinded for practical and ethical reasons, and we did not investigate the separate effects of hypoxia and cognitive training. However, the present findings are supported by our parallel four-arm double-blinded trial in healthy participants [33] and neuropsychological performance is generally not affected by placebo effects [41]. Lastly, the primary LMM analysis assumes missingness at random; because attrition was greater in H-CT than TAU, results should be interpreted with this assumption in mind.

In conclusion, although there was no effect of the intervention on the primary broad cognitive outcome, findings from the study suggest that H-CT improves executive functions and enhances daily functioning and mood. At the mechanistic level, the intervention modulated presynaptic [^11^C]UCB-J binding in the nucleus accumbens, which correlated with executive function improvement, and produced long-term changes in working memory-related dPFC and occipital activity. The study thus provides important contributions to the emerging evidence that moderate hypoxia exposure in combination with cognitive training may be a promising pro-cognitive treatment for psychiatric populations marked by cognitive impairment [15]. Future studies should include longer follow-up times for improved mechanistic insight and determine optimal treatment regimens feasible for clinical implementation.

## Supporting information

CONSORT checklist

## Data Availability

Deidentified data is available from the corresponding author upon reasonable request.

## Acknowledgements

We would like to express our gratitude to all participants for generously contributing their time and effort to the study. We also extend our thanks to medical students Ida Plougmann Østergaard and Katrine Krabbe Thommesen for their valuable contributions during their research internships, as well as to the research assistants at the NEAD Centre for their work with neuropsychological testing, fMRI scanning, and treatment monitoring. The technical assistance of Nadine Becker von-Buch is also gratefully acknowledged. This study was funded by the European Research Council (ERC) Consolidator Grant awarded to PI Kamilla Miskowiak (grant no. 101043416). In addition, the study has received financial support from the AP Møller foundation (grant no. 2022-00134). Medical students, Anne Bügel Fisker Madsen and Barbara Emilie Rotbøl Ørum, were supported by six-month research scholarships from the Lundbeck Foundation (grant no. R432-2023-94). The funding bodies had no role in the data collection, analysis, or interpretation of data, in the writing of the article, or in the decision to submit the article for publication.

## Author Contributions

Conceptualization: KWM, HE. Data curation: JMS, VD, KC, ABFM, BERØ, CFB, MBJ, LVK. Methodology: JMS, VD, AJ, JM, PPS, CS, GMK, MBJ, LVK, HE, KWM. Formal analysis: JMS, VD, AJ, JM, MM, CS, KWM. Writing–original draft: JMS, VD, KWM. Writing–review and editing: All authors. JMS and VD assessed and verified the underlying data. All authors had full access to all the data in the study and had final responsibility for the decision to submit for publication.

