## Supplementary material for "Moderate hypoxia and cognitive training for cognitive impairment in mood disorders: a randomized controlled trial"

### Supplementary Materials

### Supplementary Methods

#### In- and exclusion criteria

Eligibility criteria were 18-65 years of age, fluency in Danish, diagnosis with bipolar disorder (BD) or major depressive disorder (MDD) confirmed using the Schedules for Clinical Assessment in Neuropsychiatry (SCAN) [1], full or partial remission from mood symptoms (defined as scores ≤14 on the Hamilton Depression Rating Scale-17 items version (HDRS-17) [2] and Young Mania Rating Scale (YMRS) [3]) and objective and/or subjective cognitive impairment. Cognitive impairment was assessed with the objectively verified screening tool Screen for Cognitive Impairment in Psychiatry (SCIP; defined as total score or two subscales ≥0.5 standard deviations (SD) below expected score) [4, 5] and the questionnaire Cognitive Complaints in Bipolar Disorder Rating Assessment (COBRA; defined as total score ≥14 [6, 7]). Sex was assessed through the Danish civil registration number system (CPR). Ethnicity data was not collected. Individuals were excluded if they presented with schizophrenia spectrum disorders, organic mental disorders, history of neurological disorder or severe head trauma, alcohol or substance abuse, daily use of ≥22.5 mg oxazepam, dyslexia, or ECT <3 months prior to inclusion. For safety reasons, individuals with previous altitude sickness, significant medical conditions (e.g., heart disease, lung disease, diabetes), own or first-degree family history of thromboembolic events, pregnancy, smoking, or BMI >30 could not participate. Participants were excluded from functional magnetic resonance imaging (fMRI) assessments if they had claustrophobia or MRI incompatible metal implants. Participants were not eligible for the positron emission tomography (PET) scan if they had significant occupational exposure to radioactivity, had participated in experiments with radioactivity (>10 mSv) over the past year, or took incompatible medication (i.e., synaptic vesicle glycoprotein 2A (SV2A) binding agents).

#### Healthy participants

We have conducted a parallel double-blinded study in healthy individuals without psychiatric history aged 18-50 with the same exclusion criteria [8]. Here, participants were randomized and completed three weeks of hypoxia (*n*=60) or normoxia (*n*=54) with cognitive training or sham training for 3.5 hours, six days per week (18 sessions in total) but otherwise followed the same study protocol as the present study [9]. We included data from these participants for analyses on changes in routine blood values throughout the intervention period (inclusion, day 8, day 19, and one-month follow-up). These healthy participants were younger (median age=25) than the included individuals with mood disorders (median age=35), and we therefore included *n*=34 age, sex, and IQ-matched healthy controls (HC) for cognitive test score standardization for the present analyses. These HC had been recruited through websites or blood banks in the Capital Region of Denmark for our recent trial on cognitive remediation in virtual reality (VR) (NCT06038955) [10, 11]. They had no personal history of treatment-requiring psychiatric illness, and were excluded for dyslexia, alcohol or substance abuse, significant medical conditions, and history of severe head trauma.

#### Description of treatment sessions

The intervention took place in a sealed 20 $m^2$ treatment room at the Department of Psychology, University of Copenhagen, Denmark that was specially built for simulated altitude training. The room had four desks with dividers between them and treatment groups consisted of 2-4 participants at a time. We chose the group intervention format for practical reasons, as this allowed us to treat up to four participants at the same time. Fresh air with reduced $O_2$-levels was blown into the room by a 4 kW air compressor with a safety-approved system developed by Höhenbalance, Austria. Participants entered the room at 16% $O_2$ (≈2,200m altitude), and oxygen levels were then reduced to 12% (≈4,400m altitude). On the first day of treatment, this occurred over two hours to allow for acclimatization. For the following days, $O_2$ reduction lasted a mean 47 ± 5 minutes. Participants always wore a pulse oximeter (Shanghai Berry Electronic Tech Co., Ltd, Shanghai, China) which measured pulse and blood oxygen saturation ($SpO_2$). Values were transmitted to tablets outside the room via Bluetooth, which were continuously monitored by unblinded research personnel to ensure participant safety. For the first day and for the first 30 min of a treatment session on subsequent days (i.e., during acclimatization), participants relaxed by their desks. They then completed cognitive training on the Danish version of the HappyNeuron Pro online platform (France, www.happyneuronpro.com) for 3x40 minutes. The HappyNeuron program involved 26 training tasks that target processing speed, verbal and visual learning, working memory, and executive functions. Each task has 30 difficulty levels, and participants automatically advanced to a more challenging level once they achieved an 80% success rate for three consecutive trials. In this way, the program employed principles of neuroplasticity-based learning by being neuroadaptive, intensive, and rewarding. This was interleaved with breaks where participants relaxed for 10 minutes and used an in-room treadmill for 10 minutes at low speed (max 3km/h) to counter adverse effects of prolonged sedentary activity on cognition. Participants had a short bathroom break halfway through the session and time spent outside the room was recorded. Water, coffee, tea, and snacks were available during treatment sessions. Before entering the treatment room and after treatment completion, participants were interviewed about their well-being and sleep quality and completed a Visual Analogue Scale (VAS) about their subjective state and mood. They also completed the Environmental Symptoms Questionnaire (ESQ) [12] after each treatment session to systematically record any cerebral symptoms of altitude sickness. There was always unblinded study personnel present during treatment sessions who continuously monitored safety and who the participants could come in contact with at any time (KC, ABFM, BERØ, VD, and JMS). Medical doctors (CBF, MBJ, and LVK) were available by phone in the event of medical emergencies. In the beginning of the second week of treatment, participants had their mood symptoms systematically rated with HDRS-17 and YMRS to ensure mood stability. In the event of increased symptomatology (scores on either scale >10), we repeated the ratings in the beginning of the third week. In case of serious elevation of mood symptoms at any timepoint, the primary investigator (KWM, professor of Psychology) was consulted, and exclusion from the study was considered.

#### Assessment of treatment adherence and safety

To assess treatment adherence, we monitored number of completed treatment sessions, time spent outside the treatment room during each session, and time spent as well as distance walked on the treadmill for each participant in the hypoxia with cognitive training (H-CT) group. We also calculated the total hours spent on cognitive training and assessed the maximum achieved levels on the training tasks that a participant had completed on a given day using data directly from the HappyNeuron program. We investigated descriptive statistics (means and SDs for normally-distributed variables, medians and IQR for skewed data) of these adherence measures to assess average treatment dose.

For treatment safety, we recorded the average $SpO_2$ and pulse for each day using data from the oximeters. We also collected total ESQ scores, subjective sleep quality and tiredness, and change in physical wellbeing and all VAS measures from each session. These values were reported as means and SD or medians and IQR, depending on data distribution, to assess treatment tolerability. We also compared these variables between participants in the H-CT group who completed and who did not complete the intervention, respectively, using independent samples *t*-tests for normally-distributed data and Mann Whitney *U*-tests for skewed data.

#### Outcome standardization and domain calculation

We standardized behavioral scores based on the age-, sex- and IQ-matched HC sample. For rating/questionnaire data on functioning, quality of life, subjective cognition, and sleep quality, mean percent blood-oxygen-level-dependent (BOLD) signal change, and research blood sample values (erythropoietin (EPO), brain-derived neurotrophic factor (BDNF), and vascular endothelial growth factor (VEGF)) we standardized scores based on the H-CT and treatment as usual (TAU) groups' baseline scores. Relevant variables were inversed so higher scores always corresponded to better performance/functioning.

Tertiary cognitive outcomes were domain scores from the full neuropsychological test battery applied at baseline, treatment completion and one-month follow-up (and following treatment completion for TAU group participants who underwent H-CT after their one-month follow-up visit). The full battery included tests grouped within the following cognitive domains: Processing speed: The Repeatable Battery for the Assessment of Neuropsychological Status (RBANS) Coding and Trail Making Test (TMT)-A [13, 14]; Attention: RBANS Digit Span [14], and Rapid Visual Processing (RVP) accuracy, mean latency, total false alarms, and probability of hit (CANTAB, Cambridge Cognition Ltd.); Working memory: Wechsler Adult Intelligence Scale (WAIS)-III Letter-Number Sequencing (LNS) [15] and Spatial Working Memory (SWM) between errors and strategy (CANTAB); Verbal learning and memory: The Rey Auditory Verbal Learning Test (RAVLT) total recall list I-V, immediate recall, and delayed recall [16]; Executive functions: TMT-B [13], Verbal Fluency letters "S" and "D" [17], One-Touch Stockings of Cambridge (OTS) mean choices to correct, problems solved on first choice, and mean latency to correct (CANTAB), and Wisconsin Card Sorting Task (WCST) perseverative errors [18]; and Facial expression recognition: Emotion Recognition Test (ERT) average hit rate across emotions and overall reaction time (CANTAB). To mitigate effects of repeated testing, parallel versions of the RAVLT and RBANS were used in a counter-balanced order for each assessment point.

The tertiary domain scores were calculated by averaging relevant *z*-scores. As OTS 'mean choices to correct' was secondary outcome, this was not included in the executive functions domain calculations. We also computed a global cognition composite by averaging processing speed, attention, working memory, verbal learning and memory, and executive functions domain scores.

Tertiary rating-based and self-reported outcome domains were evaluated at all assessment points and included the following measures: Quality of life: World Health Organization Quality of Life (WHOQOL-BREF) [19] and Assessment of Quality of Life (AQoL) [20]; Subjective cognition: Cognitive Complaints in Bipolar Disorder Rating Assessment (COBRA) [7]; Functioning: Functional Assessment Short Test (FAST) (baseline and one-month follow-up only) [21], Work and Social Adjustment Scale (WSAS) [22], and Sheehan Disability Scale (SDS) [23]; and Sleep quality: Pittsburgh Sleep Quality Index (PSQI) [24]. Tertiary domain scores were calculated by averaging relevant *z*-scores. The functioning outcome Cognitive Assessment in Virtual Reality (CAVIR) [25] composite score was a secondary outcome and was therefore not included in the functioning domain calculation. Finally, we added HDRS-17 and YMRS total raw scores to compute a total mood symptom score for post-hoc mood-corrected analyses.

#### **PET analysis**

***PET power calculation***

The power calculation for the positron emission tomography (PET) imaging analyses was based on two assumptions: (1) the mean non-displaceable binding potential ($BP_{ND}$) in the frontal cortex is $M$±SD=3.36±0.38, based on in-house data for psychiatrically healthy individuals, and (2) H-CT vs.

TAU will lead to a 10% increase in $BP_{ND}$ in the frontal cortex and/or hippocampus. To achieve >80% power to identify this difference in $BP_{ND}$ for H-CT vs. TAU at treatment completion at an α-level of 0.05 (two-tailed), we aimed to include a total of 40 participants (H-CT: $n$=20; TAU: $n$=20). However, as the PET scanner was temporarily unavailable during the early trial period due to technical issues, we achieved an actual sample size of 28 (H-CT: $n$=13; TAU: $n$=15) participants (see flow chart and missing data below).

***PET subsample and missing data***

See **Supplementary Figure S1** for flow diagram of the subsample included in the PET imaging analysis. Among the 64 participants that were enrolled and randomized, 55 participants (26 H-CT, 29 TAU) completed the week 4 assessments (treatment completion). Of these, 28 participants underwent and completed a PET scan in week 4 (13 H-CT, 15 TAU). Thus, 27 participants were not included in the PET imaging analysis due to the following reasons: the scanner was unavailable due to temporary closure (n=7), they were not eligible for either PET or MRI (n=4), they did not have time (n=4) or did not want to (n=2) undergo PET, they were evaluated as too psychiatrically vulnerable for PET by the research personnel (n=3), they had claustrophobia (n=2), difficult venous access (n=1), mood symptom exacerbation (n=1), issues with the tracer from the radiochemistry production (n=2), or technical issues occurred during PET acquisition (n=1). Baseline demographic characteristics were well-balanced between the treatment groups for the PET subsample, with no differences in age, sex, or verbal IQ ($ps \geq 0.29$).

***Data acquisition***

PET imaging data were acquired the Copenhagen University Hospital, Rigshospitalet, Denmark using a high-resolution research tomography (HRRT) PET scanner (CTI/Siemens, Knoxville, TN, USA).

After a six-minutes transmission scan, participants underwent a 90-minutes emission scan, which started at the time of the intravenous bolus injection of between 100 and 510 MBq [$^{11}$C]UCB-J administered over ~20 seconds. PET data were obtained in 3D list mode and reconstructed into a total of 37 frames (8x15 s, 8x30 s, 4x60 s, 5x2 min, 10x5 min, and 2x10 min) using a 3D OP-OSEM algorithm with modelling of the point-spread-function, and attenuation corrected using the HRRT maximum a posteriori transmission reconstruction method (MAP-TR). Each image frame consisted of 207 planes of 256×256 voxels of 1.22×1.22×1.22 mm$^3$.

***PET pre-processing***

Motion correction was applied to all PET images using the Automated Image Registration (AIR) software with the reconcile command (v. 5.2.5) [26]. The rest of the image pre-processing was conducted using the PVElab pipeline (Neurobiology Research Unit, Copenhagen; https://nru.dk/pveout/). PVElab used an unfiltered summation PET image that was automatically co-registered to the participant's T1-weighted MR image from their follow-up scan using SPM12. Multispectral segmentation (i.e., both T1 and T2-weighted MR images) was then applied to extract tissue time-activity curves from each automatically defined region of interest (ROI) [27–29]. We visually inspected all planes to confirm accurate co-registration and ROI placement. No manual correction was needed. No partial volume effects correction was applied. The whiter matter (centrum semiovale) reference region was acquired from the PVElab region and was further eroded twice with a 3D erosion operator to reduce partial volume effects - the final volume was $M\pm SD$=7.45±2.63 mL.

***Kinetic modelling***

Tissue time-activity curves were fitted to a simplified reference tissue model 2 (SRTM2) to estimate $BP_{ND}$ using the centrum semiovale as reference [30, 31]. We used a population based k2' value of

0.035 min-1 for each fit, which was based on the median k2 value estimated from one-tissue-compartment-models from our prior [$^{11}$C]UCB-J PET study in psychiatrically healthy individuals [31].

***PET regions of interest***

Mechanistic PET outcomes were differences in [$^{11}$C]UCB-J $BP_{ND}$ (as a readout of presynaptic density) between H-CT vs. TAU at treatment completion (week 4). Although the PET outcomes were mechanistic and thus exploratory, our a priori selected ROIs were the hippocampus and frontal cortex – two key regions linked to decreased neuroplasticity and cognitive deficits in mood disorders. To explore possible treatment group differences in presynaptic density outside these regions, we also compared average $BP_{ND}$ between H-CT and TAU for the following six subcortical ROIs: amygdala, nucleus accumbens, caudate nucleus, putamen, thalamus, striatum, and 23 cortical ROIs: neocortex, insula, anterior cingulate cortex, posterior cingulate cortex, middle frontal gyrus, precentral gyrus, postcentral gyrus, gyrus rectus, orbitofrontal gyri, inferior frontal gyrus, superior frontal gyrus, anterior temporal lobe (medial part), anterior temporal lobe (lateral part), posterior temporal lobe, parahippocampal and ambient gyri, superior temporal gyrus, middle and inferior temporal gyri, fusiform gyrus, superior parietal gyrus, inferolateral remainder of parietal lobe, lingual gyrus, cuneus, and lateral remainder of occipital lobe. All ROIs were bilateral. The neocortex was defined as the weighted average of the individual subregions: frontal, parietal, temporal, occipital, and insular cortices.

#### Functional magnetic resonance imaging

##### *Spatial working memory N-back paradigm*

The fMRI paradigm was a spatial working memory N-back task programmed in E-prime 2.0 (Psychological Software Tools, Pennsylvania, USA). Stimuli were shown on a screen that participants viewed through an angled mirror in the scanner. Participants were shown a 5x5 grid where a yellow circle appeared for 300 ms followed by an empty grid for 1200 ms. They were told to indicate with a button press whenever the yellow circle appeared in the same grid square as the one from *N* (1 or 2) steps back (i.e., 1-back and 2-back conditions). In a sensorimotor control task (0-back), they had to respond when the yellow circle appeared in any of the grid corners. The blocks had an average of three target trials and were presented successively five times interleaved with 8 s fixation crosses (15 blocks in total, total task length=7 min 35 s).

##### *fMRI data acquisition*

Functional magnetic resonance imaging data were collected at the University Hospital of Copenhagen, Denmark with a 3 Tesla SIEMENS Prisma scanner, using a 32-channel head-neck coil. Functional BOLD T2*2-weighted images were acquired with a multi-band multi-echo (MB3ME3) sequence (parameters: TR=1300 ms, TEs=15, 34, and 54 ms, flip angle=70°, matrix size=76x76, slices per volume=48, slice thickness=2.5 mm, voxel size=2.5 mm isotropic, and a 3x multiband acceleration factor). We acquired 228 volumes in total for the spatial N-back fMRI task. In addition to T2-weighted images, we acquired structural T1-weighted images for registration of functional images to the standard Montreal Neurological Institute (MNI) template. T1-images were collected with a standard Magnetization Prepared Rapid Gradient Echo (MPRAGE) pulse sequence (parameters: TR=2000 ms; TE=2.58 ms; flip angle=8°; matrix size=256x256; slices per volume=224; slice thickness=0.9 mm, and voxel size=0.9 mm isotropic). Finally, we collected fieldmap data to correct for geometric distortions arising from $B_0$ inhomogeneities with a gradient echo sequence (TR=444 ms; TE1=4.92 ms; TE2=7.38 ms; flip angle=60°). Fieldmap images were computed based on the phase difference between two echoes with the FMRIB Software Library (FSL) version 6.0.5.2 [32].

***Pre-processing pipeline***

We used the fMRIPrep 25.1.1 software package to preprocess the fMRI data [33]. This included generating a BOLD reference volume by aligning and averaged three single-band references, estimating head-motion parameters according to the BOLD reference, susceptibility distortion correction based on the fieldmap, slice-time correction, combining the multi-echo time series, co-registration to the T-weighted image, resampling to the standard MNI152Lin6Asym space with 2x2x2 voxel size, spatial smoothing with a 5 mm full-width-half-maximum gaussian kernel, and, finally, high-pass temporal filtering with a 100 s cut-off. We visually expected every participant's MNI template registration as well as head movement parameter plots to ensure the quality of the pre-processing.

***Subject-level analysis***

We modelled the spatial N-back fMRI task at the subject-level in the FMRI Expert Analysis Tool (FEAT). The events were convolved with a double-gamma hemodynamic response function. In the subject-level general linear model (GLM), we applied motion correction by including the six motion parameters estimated during motion correction as nuisance regressors. We then calculated framewise displacement (FD) as the sum of the absolute values of the derivative of the six motion parameters [34]. Volumes with an FD>0.5 mm were removed, and we generated additional nuisance regressors for the high-motion volumes and included these in the subject-level analysis to account for motionrelated artifacts through volume censoring (spike regression). We excluded scans from the subsequent group-level analyses if the number of removed volumes exceeded 25% (=57 out of 228 volumes). We also excluded participants with mean FD>0.3 mm across the paradigm.

***Statistical analysis of in-scanner behavioral data***

We recorded in-scanner behavioral performance during the spatial N-back task for correlations with BOLD response. The outcome of interest was discriminability index (*d*') which is a measure of response accuracy during the different conditions of the paradigm (i.e., 0-, 1-, and 2-back), respectively. The *d*' values were calculated with the following equation: ((no. of hits+0.5)/(total no. of targets+1))-((no. of false alarms+0.5)/(no. of distractors+1)). This calculation yielded scores ranging from 0 to 1 with scores closer to 1 indicating better accuracy, which were *z*-transformed according to H-CT and TAU groups baseline data. We investigated Pearson's correlations between mean percent signal change in clusters showing significant time*treatment interaction effects and the *d*' of the 1-back condition for the 1>0-back contrasts and the combined *d*' of the 1- and 2-back conditions for the 2>0-back- and 2>1-back contrasts, respectively.

***fMRI subsample and missing data***

Two participants (one H-CT, one TAU) dropped out of the study before baseline assessments and were therefore not scanned. Four participants (two H-CT, two TAU) were excluded from fMRI assessments because of risk of metal implants or claustrophobia. Three participants (all H-CT) were scanned at baseline only, but data was not included in the analyses because of large artefacts. Three baseline scans (two H-CT, one TAU) were excluded from analyses because of excessive movement. This yielded a baseline sample of *n*=26 H-CT and *n*=26 TAU. Five H-CT dropped out and were not scanned at follow-up. One participant (TAU) did not complete the spatial N-back task during follow-up and was therefore excluded from analyses. Two follow-up scans (both H-CT) were excluded because of excessive movement. Hence, the sample size at follow-up was *n*=21 H-CT and *n*=26 TAU. We included data from all applicable scans in the LMMs of mean percent signal change in our right dorsolateral prefrontal cortex (dlPFC) region of interest (ROI). In the dorsal prefrontal cortex (dPFC) volume of interest (VOI) and whole brain analyses, we only included participants with complete baseline and follow-up data (*n*=19 H-CT; *n*=25 TAU).

#### Comparisons between treatment groups and with healthy control sample

We computed summary statistics on demographic and clinical variables for the participants in the H-CT and TAU groups, respectively. We calculated means and SD for continuous, normally distributed data, medians and interquartile range (IQR) for continuous, skewed data, and counts and percentages for categorical data. In accordance with CONSORT reporting guidelines, we did not perform inferential statistical comparisons between treatment groups. However, we tested for differences in demographic variables and outcome data between the *n*=64 study participants and the *n*=34 HC included for standardization of neuropsychological test performance. Here, we used *t*-tests for comparisons of continuous, normally distributed variables, Mann-Whitney *U* tests for comparisons of continuous, skewed variables, and Chi-Square tests for comparisons of categorical data.

#### Statistical analyses of treatment as usual group

We performed exploratory analyses in the subsample of the TAU group who completed the intervention following their waitlist condition. Here, we investigated change in primary, secondary, and tertiary outcomes from the one-month follow-up timepoint (i.e., before commencing H-CT treatment) to after treatment completion using LMMs to explore potential treatment effects. We used an unstructured covariance pattern, and fixed effects were time and stratum.

### Supplementary Results

#### Comparisons with healthy control sample

The study participants with mood disorders (*n*=64) were comparable in age, sex, and IQ to the HC included for standardization of cognitive outcomes (*n*=34) (*p*s≥0.12). However, a larger proportion of the HC were currently employed or studying ($p<0.001$). As expected, the study participants had more subsyndromal depressive symptoms ($p<0.001$) but did not show elevated levels of (hypo)manic symptoms compared to HC ($p=0.47$). The study participants had lower scores on all cognitive outcomes (*p*s≤0.03) except processing speed ($p=0.06$), verbal memory ($p=0.38$) and facial emotion recognition ($p=0.32$). They also reported more subjective cognitive complaints ($p<0.001$) and showed poorer functioning on CAVIR ($p=0.004$) and FAST ($p<0.001$). See **Supplementary Table S1** for all group comparisons.

#### Treatment adherence and dose

The *n*=26 participants from the H-CT group who completed the intervention attended a median of 15 (IQR=15-15.8) treatment sessions. Six participants included in the beginning of the trial period completed between 16-18 treatment sessions (i.e., more sessions than defined as 100% treatment completion in the protocol). We chose to adjust the treatment schedule from 18 to 16 days of treatment early in the trial as we found that most participants needed to have two days rather than one day off per week during the three-week intervention period. Each day, participants spent a median of 1 minute (IQR=0.6-2 minutes) outside the room in addition to their scheduled restroom break in the middle of the session. They spent 27±4 hours in total on cognitive training and reached an average of level 13±2 out of 30 on the day where they achieved their highest cognitive levels. They walked a median 25,

IQR=23-27 minutes per day, corresponding to approximately 1.2 km of slow-paced treadmill walking. See **Supplementary Table S6** for a full overview of treatment adherence and dose.

#### Treatment tolerability

The H-CT intervention led to the expected drop in $SpO_2$ levels with median blood oxygen saturations of 85%, IQR=83-87%. Average pulse was 84±9 beats per minute. Participants in the treatment group who completed the intervention had low degrees of cerebral side-effects. Accordingly, the median ESQ total score was 4, IQR=1-5 (possible scores ranging from 0 to 55). Participants generally reported good sleep quality during the intervention (median=8.5, IQR=7-9 out of 10, where 10 corresponded to as good sleep quality as usual) and reported minimal changes in wellbeing or subjective state from before to after intervention as measured with interviews and VAS scales (0 to 10 scales, mean change scores ranging from -0.3 to 0.7 across measures). **See Supplementary Table S7** for a full overview of treatment safety outcomes.

##### *Completers vs. non-completers*

The participants assigned to H-CT who did not complete the intervention ($n$=8) reported higher average ESQ scores than participants who completed the intervention ($n$=26) (median scores=12, IQR=7-13 vs 4, IQR=1-5, $p$=0.02). Non-completers reported more dizziness and nausea after the sessions compared to before treatment than completers on a VAS ($p$s≤0.03). There were no differences in other safety parameters between groups ($SpO_2$, pulse, sleep quality, tiredness, or change in wellbeing, happiness, sadness, vigilance, or anxiety, $p$s≥0.09). See **Supplementary Table S7** for a full overview of comparisons of treatment safety outcomes between completers and non-completers in the H-CT group.

#### Routine blood parameters

We compared changes in routine blood parameters (hemoglobin, erythrocytes, reticulocytes, and thrombocytes) in participants who completed the intervention with LMMs. We investigated differential change in the H-CT group in these routine blood values with the normoxia group from our parallel study in healthy participants (N-HP) [9] and the TAU group, respectively, as reference group. The N-HP group was included for comparisons as we did not take routine blood samples in the TAU group on day 8. The models used an unstructured covariance pattern, baseline constraints, and had time, treatment, and time*treatment interaction as fixed effects. Linear mixed models showed no differences in blood values from baseline (inclusion) to day 8, day 19, or one-month follow-up between H-CT and N-HP groups ($p$s≥0.12) or between H-CT and TAU groups ($p$s≥0.18) (Figure S2). In contrast, the hypoxia group in healthy individuals (H-HP) had elevated blood values (all within reference values) compared to N-HP for hemoglobin on day 8 ($p$=0.01) and day 19 ($p$<0.001), erythrocytes on day 19 ($p$=0.02), reticulocytes on day 8 ($p$=0.008), and thrombocytes on day 8 ($p$=0.04) and day 19 ($p$<0.001), which had all normalized at one-month follow-up ($p$s≥0.34). See **Supplementary Table S8** for raw values for all groups across the four timepoints.

#### Additional mechanistic analyses

***Post hoc correlations between change in cognition and change in functioning***

As the H-CT group had improved executive functions (secondary outcome and tertiary domain score) as well as improved functioning, subjective cognition, and mood after the intervention, we performed exploratory correlational analyses between change in executive functions and change in functioning. Pearson's correlation analyses showed no significant correlations between change in OTS 'mean choices to correct' or executive functions domain scores and change in functioning, subjective cognition scores, or mood from baseline to treatment completion or baseline to one-month follow-up, respectively ($p$s≥0.17) (**Supplementary Table S2**).

***Post hoc correlations between change in secondary outcome and treatment dose***

We explored if treatment dose was associated with improvements in the secondary cognitive outcome with correlation analyses within the H-CT group. There were no significant Pearson's correlations between change in OTS 'mean choices to correct' from baseline to treatment completion or baseline to one-month follow-up and number of treatment sessions, total hours spent with HappyNeuron training, or max level achieved in HappyNeuron ($p$s≥0.12) (**Supplementary Table S3**).

***Analyses in treatment as usual group***

See **Supplementary Table S5** for results of the exploratory analyses in the TAU group. In the $n$=18 TAU participants who subsequently completed the H-CT intervention, there was no change in the primary outcome 'speed of complex cognitive processing' from before treatment to end of treatment ($p$=0.11). However, scores on the secondary functioning outcome (CAVIR global composite) (treatment effect=0.45, $p$=0.02) improved from before to after treatment, which did not occur in the primary analyses of H-CT vs. TAU. Also contrasting the primary analyses, there was no improvement in the secondary cognitive outcome OTS 'mean choices to correct' ($p$=0.13) from before to after treatment. On the tertiary domains, the TAU group improved after treatment on working memory (treatment effect=0.35, $p$=0.03), verbal learning and memory (treatment effect=0.60, $p$=0.02), and the global cognition composite (treatment effect=0.30, $p$=0.01), all of which were non-significant in the main analyses. All other domains, including executive functions, facial emotion recognition, quality of life, subjective cognition, and functioning, which showed effects of H-CT in the primary analyses, were non-significant from before to after treatment in the TAU group ($p$s≥0.08).

#### Supplementary Tables

**Supplementary Table S1.** Group comparisons between study participants and healthy controls included for standardization of outcome measures.

| Measure | Mood disorders | Healthy controls | *p*-value |
|---|---|---|---|
| N | 64 | 34 | - |
| Age, median (IQR) | 35.0 (28.8, 53.3) | 32.5 (27.0, 46.8) | 0.12 |
| Sex, no. female (%) | 44 (69%) | 19 (56%) | 0.30 |
| Years of education, mean (SD) | 15.8 (2.5) | 16.6 (2.0) | 0.10 |
| Verbal IQ, mean (SD) | 112.2 (6.3) | 113.4 (5.4) | 0.35 |
| Currently employed/studying, no. yes (%) | 44 (69%) | 31 (91%) | **<0.001** |
| HDRS-17, median (IQR) | 3.0 (1.0, 6.0) | 0.0 (0.0, 1.0) | **<0.001** |
| YMRS, median (IQR) | 0.0 (0.0, 1.0) | 0.0 (0.0, 0.8) | 0.47 |
| Speed of complex cognitive processing, mean (SD) | -0.5 (1.0) | 0.0 (0.5) | **0.007** |
| OTS Mean choices to correct, mean (SD) | -1.1 (1.6) | 0.0 (1.0) | **<0.001** |
| CAVIR global composite, mean (SD) | -0.6 (1.0) | 0.0 (0.6) | **0.004** |
| Processing speed, mean (SD) | -0.4 (1.0) | 0.0 (0.8) | 0.06 |
| Attention, mean (SD) | -0.8 (2.0) | 0.0 (0.6) | **0.03** |
| Working memory, mean (SD) | -0.8 (1.1) | 0.0 (0.7) | **<0.001** |
| Verbal memory, mean (SD) | -0.2 (1.4) | 0.0 (0.9) | 0.38 |
| Executive functions, mean (SD) | -0.5 (0.8) | 0.0 (0.4) | **<0.001** |
| Global cognition, mean (SD) | -0.5 (0.9) | 0.0 (0.4) | **0.002** |
| Facial emotion recognition, mean (SD) | 0.2 (0.9) | 0.0 (0.7) | 0.32 |
| COBRA, median (IQR) | 23.0 (18.0, 27.3) | 10.0 (6.0, 12.0) | **<0.001** |
| FAST, median (IQR) | 16.5 (11.0, 24.5) | 1.0 (0.0, 2.0) | **<0.001** |
| *Mood disorder diagnosis, no. (%)* | | | |
| BD | 28 (44%) | - | - |
| MDD | 36 (56%) | - | - |
| *BD type, no. (%)* | | | |
| 1 | 10 (36%) | - | - |
| 2 | 18 (64%) | - | - |
| Illness duration (years), mean (SD) | 20.9 (13.7) | - | - |
| Depressive episodes, median (IQR) | 5.5 (3.0, 13.0) | - | - |
| Hypomanic episodes, median (IQR) | 0.0 (0.0, 3.5) | - | - |
| Manic episodes, median (IQR) | 0.0 (0.0, 0.0) | - | - |
| Psychiatric hospitalizations, median (IQR) | 0.0 (0.0, 1.3) | - | - |
| *Psychiatric treatment, no (%)* | | | |

| | | | |
|---|---|---|---|
| None | 12 (19%) | - | - |
| GP | 20 (32%) | - | - |
| Private psychiatrist | 16 (25%) | - | - |
| Mental health services | 15 (24%) | - | - |
| Antidepressants, no. yes (%) | 28 (44%) | - | - |
| Quetiapin stabilization, no. yes (%) | 5 (7.8%) | - | - |
| Lamotrigin, no. yes (%) | 17 (27%) | - | - |
| Lithium, no. yes (%) | 17 (27%) | - | - |
| ADHD medication, no. yes (%) | 4 (6.3%) | - | - |
| PN psychotropic medication, no. yes (%) | 26 (41%) | - | - |
| No psychotropic medications, no. yes (%) | 15 (23%) | - | - |
| Somatic medications, no. yes (%) | 19 (30%) | - | - |

**Note:** Table shows group comparisons between study participants with mood disorders and healthy controls included for standardization of neuropsychological test performance and neuroimaging baseline comparisons as well as clinical characteristics for the full mood disorder sample. Means, SD, and independent samples t-tests for continuous, normally-distributed data; medians, IQR, and Mann-Whitney U tests for continuous, skewed data; counts, percentages, and Chi-Square tests for categorical data. **Missing data**: IQ: Two MD. Speed of complex cognitive processing: Two MD. OTS mean choices to correct: Two MD. CAVIR global composite: Two MD. Processing speed: Two MD. Attention: Two MD. Working memory: Two MD. Verbal memory: Two MD. Executive functions: Two MD. Global cognition: Two MD. Facial emotion recognition: Six MD. COBRA: Four MD. FAST: Two MD. Hypomanic episodes: One MD. Psychiatric treatment: One MD. **Abbreviations**: ADHD=Attention Deficit Hyperactivity Disorder; BD=Bipolar disorder; CAVIR=Cognitive Assessment in Virtual Reality; COBRA=Cognitive Complaints in Bipolar Disorder Rating Assessment; FAST=Functional Assessment Short Test; GP=General practitioner; HDRS-17=Hamilton Depression Rating Scale 17 items version; IQ=Intelligence quotient; IQR=Interquartile range; MD=Mood disorders; MDD=Major depressive disorder; OTS=One-touch Stockings of Cambridge (CANTAB); PN=Per necessitate; SD=Standard deviation; YMRS=Young Mania Rating Scale.

**Supplementary Table S2.** Treatment adherence and dose for participants in the hypoxia with cognitive training group who completed the intervention.

| **Measure** | **Completed hypoxia cognitive training** |
|---|---|
| N | 26 |
| Number of treatment sessions, median (IQR) | 15.0 (15.0, 15.8) |
| Time spent outside treatment room pr session, median (IQR) | 1.1 (0.6, 2.1) |
| Total hours spent on cognitive training, mean (SD) | 27.3 (4.0) |
| Average max level acheived in cognitive training, mean (SD) | 12.8 (2.3) |
| Time spent on treadmill pr session, median (IQR) | 25.1 (23.0, 26.9) |
| Distance (km) walked on treadmill pr session, median (IQR) | 1.2 (1.0, 1.3) |

**Note:** Table shows descriptive statistics for participants in the hypoxia with cognitive training group who completed the intervention. Means and SD for continuous, normally-distributed data; medians and IQR for continuous, skewed data; counts and percentages for categorical data. **Abbreviations:** IQR=Interquartile range; Km=Kilometers. SD=Standard deviation.

**Supplementary Table S3.** Treatment safety and tolerability measures for participants in the hypoxia with cognitive training group who completed the treatment and who did not complete the treatment.

| **Measure** | **Completed hypoxia cognitive training** | **Did not complete hypoxia cognitive training** | ***p*-value** |
|---|---|---|---|
| N | 26 | 8 | - |
| Average SpO2 per session, median (IQR) | 84.6 (82.6, 87.1) | 85.0 (84.0, 86.2) | 0.87 |
| Average pulse per session, mean (SD) | 83.8 (9.3) | 76.6 (7.1) | 0.09 |
| Average ESQ total score per session (range = 0 to 55), median (IQR) | 3.5 (1.4, 5.1) | 11.5 (6.9, 13.4) | **0.02** |
| Subjective sleep quality (range = 0 to 10), median (IQR) | 8.5 (7.4, 9.1) | 7.9 (7.5, 8.2) | 0.53 |
| Subjective tiredness after each session (range = 0 to 5), median (IQR) | 2.1 (1.8, 3.0) | 2.5 (2.0, 3.4) | 0.37 |
| Change in physical wellbeing after each session (range = -10 to 10), mean (SD) | -0.2 (0.3) | -0.4 (1.0) | 0.27 |
| VAS change in happiness after each session (range = -10 to 10), mean (SD) | -0.1 (0.3) | -0.1 (0.8) | 0.84 |
| VAS change in wellbeing after each session (range = -10 to 10), mean (SD) | -0.2 (0.5) | -0.7 (1.1) | 0.14 |
| VAS change in sadness after each session (range = -10 to 10), mean (SD) | 0.2 (0.5) | 0.0 (0.9) | 0.52 |
| VAS change in vigilance after each session (range = -10 to 10), mean (SD) | -0.3 (0.7) | -0.5 (1.3) | 0.68 |
| VAS change in anxiety after each session (range = -10 to 10), mean (SD) | -0.2 (0.4) | -0.4 (0.8) | 0.28 |
| VAS change in dizziness after each session (range = -10 to 10), mean (SD) | 0.7 (0.9) | 1.7 (1.5) | **0.03** |
| VAS change in nausea after each session (range = -10 to 10), mean (SD) | 0.1 (0.4) | 1.2 (1.2) | **<0.001** |

**Note:** Table shows treatment safety parameters and comparisons between participants in the H-CT group who completed and who did not complete the intervention. Means, SD, and independent samples t-tests for continuous, normally-distributed data; medians, IQR, and Mann-Whitney U tests for continuous, skewed data; counts, percentages. **Missing data:** Two non-completers did not begin the intervention and therefore has missing data across all variables. Subjective sleep quality: Nine participants (five completers, four non-completers). **Abbreviations:** ESQ=Environmental Symptoms Questionnaire; IQR=Interquartile range; SD=Standard deviation; SpO2=Blood oxygen saturation; VAS=Visual Analogue Scale.

**Supplementary Table S4.** Routine hematological parameters mean raw scores.

| Measure | H-CT | TAU | Hypoxia (healthy) | Normoxia (healthy) | *p*-value |
|---|---|---|---|---|---|
| N | 26 | 29 | 60 | 54 | - |
| Hemoglobin baseline, mean (SD) | 8.5 (0.6) | 8.6 (0.8) | 8.8 (0.7) | 8.8 (0.7) | - |
| Hemoglobin day 8, mean (SD) | 8.5 (0.7) | - | 9.0 (0.8) | 8.8 (0.7) | 0.55 |
| Hemoglobin day 19, mean (SD) | 8.6 (0.7) | 8.6 (0.7) | 9.0 (0.8) | 8.7 (0.6) | 0.25 |
| Hemoglobin follow-up, mean (SD) | 8.5 (0.5) | 8.7 (0.7) | 8.9 (0.8) | 8.8 (0.6) | 0.15 |
| Erythrocytes baseline, mean (SD) | 0.4 (0.0) | 0.4 (0.0) | 0.4 (0.0) | 0.4 (0.0) | - |
| Erythrocytes day 8, mean (SD) | 0.4 (0.0) | - | 0.4 (0.0) | 0.4 (0.0) | 0.33 |
| Erythrocytes day 19, mean (SD) | 0.4 (0.0) | 0.4 (0.0) | 0.4 (0.0) | 0.4 (0.0) | 0.46 |
| Erythrocytes follow-up, mean (SD) | 0.4 (0.0) | 0.4 (0.0) | 0.4 (0.0) | 0.4 (0.0) | 0.17 |
| Reticulocytes baseline, mean (SD) | 12.3 (2.9) | 13.8 (4.9) | 13.4 (5.1) | 13.9 (4.0) | - |
| Reticulocytes day 8, mean (SD) | 15.1 (3.3) | - | 16.0 (4.9) | 14.4 (3.5) | 0.12 |
| Reticulocytes day 19, mean (SD) | 13.6 (3.1) | 14.9 (7.6) | 14.3 (4.4) | 14.1 (3.2) | 0.46 |
| Reticulocytes follow-up, mean (SD) | 12.4 (2.5) | 14.6 (3.4) | 14.5 (5.4) | 13.9 (3.9) | 0.32 |
| Thrombocytes baseline, mean (SD) | 230.1 (52.4) | 255.5 (45.0) | 246.6 (50.2) | 254.4 (61.1) | - |
| Thrombocytes day 8, mean (SD) | 255.2 (51.2) | - | 269.9 (52.4) | 266.5 (54.9) | 0.67 |
| Thrombocytes day 19, mean (SD) | 243.3 (45.3) | 251.9 (39.2) | 272.1 (51.5) | 249.8 (48.4) | 0.15 |
| Thrombocytes follow-up, mean (SD) | 259.7 (62.4) | 282.9 (42.9) | 247.1 (64.9) | 251.9 (49.1) | 0.18 |

**Note:** Table shows blood values at baseline, day 8 and 19 of the intervention, and one-month follow-up for the hypoxia with cognitive training, treatment as usual, hypoxia in healthy individuals substudy, and normoxia in healthy individuals substudy groups (participants who completed their intervention). *P*-value refers to linear mixed model comparing the hypoxia with cognitive training and normoxia (healthy individuals) groups. There were no significant differences in blood values between hypoxia cognitive training and treatment as usual groups at any timepoint (*p*s>0.18). **Missing data:** Hemoglobin: One participant at baseline (TAU), 33 participants at day 8 (five H-CT, 15 H-HP, 13 N-HP), 20 participants at day 19 (one H-CT, four TAU, six H-HP, nine N-HP), 59 at follow-up (five H-CT, 12 TAU, 22 H-HP, 20 N-HP). Erythrocytes: One participant at baseline (TAU), 33 participants at day 8 (five H-CT, 15 H-HP, 13 N-HP), 20 participants at day 19 (one H-CT, four TAU, six H-HP, nine N-HP), 59 at follow-up (five H-CT, 12 TAU, 22 H-HP, 20 N-HP). Reticulocytes: 12 participants at baseline (three TAU, three H-HP, six N-HP), 33 participants at day 8 (five H-CT, 15 H-HP, 13 N-HP), 20 participants at day 19 (one H-CT, four TAU, six H-HP, nine N-HP), 59 at follow-up (five H-CT, 12 TAU, 22 H-HP, 20 N-HP). Thrombocytes: One participant at baseline (TAU), 33 participants at day 8 (five H-CT, 15 H-HP, 13 N-HP), 20 participants at day 19 (one H-CT, four TAU, six H-HP, nine N-HP), 59 at follow-up (five H-CT, 12 TAU, 22 H-HP, 20 N-HP). **Abbreviations:** H-CT=Hypoxia with cognitive training; H-HP=Hypoxia (healthy individuals); N-HP=Normoxia (healthy individuals); SD=Standard deviation; TAU=Treatment as usual.

**Supplementary Table S5.** Correlation matrix of change in executive functions and change in self-reported/rating outcomes that showed improvement after the intervention in the hypoxia with cognitive training group.

| | **Completed hypoxia with cognitive training (*n*=26)** | | | | | | | |
|---|---|---|---|---|---|---|---|---|
| | **Treatment completion** | | **Follow-up** | | **Treatment completion** | | **Follow-up** | |
| | ΔOTS 'mean choices to correct' | | | | ΔExecutive functions | | | |
| | *r* | *p*-value | *r* | *p*-value | *r* | *p*-value | *r* | *p*-value |
| ΔFunctioning | -0.04 | 0.86 | -0.25 | 0.22 | 0.24 | 0.24 | 0.11 | 0.59 |
| ΔSubjective cognition | -0.26 | 0.19 | -0.23 | 0.27 | 0.22 | 0.28 | -0.003 | 0.99 |
| ΔMood | -0.21 | 0.31 | 0.28 | 0.17 | -0.35 | 0.08 | 0.17 | 0.42 |

**Note:** Table shows Pearson's correlation matrix of change-scores in relevant cognitive measures and self-report/rating measures. **Missing data:** Subjective cognition: Two H-CT at follow-up. Mood: One H-CT at follow-up **Abbreviations:** H-CT=Hypoxia with cognitive training; OTS=One-touch stockings of Cambridge (CANTAB).

**Supplementary Table S6.** Correlation matrix of change in secondary cognitive outcomes and treatment dose variables in the hypoxia with cognitive training group.

| | **Completed hypoxia with cognitive training (*n*=26)** | | | |
|---|---|---|---|---|
| | **Treatment completion** | | **One-month follow-up** | |
| | ΔOTS 'mean choices to correct' | | ΔOTS 'mean choices to correct' | |
| | *r* | *p*-value | *r* | *p*-value |
| Number of treatment sessions | -0.14 | 0.51 | -0.21 | 0.29 |
| Total hours spent on cognitive training | -0.15 | 0.47 | -0.15 | 0.46 |
| Average max level achieved in cognitive training | -0.31 | 0.12 | -0.31 | 0.12 |

**Note:** Table shows Pearson's correlation matrix of change-scores in the secondary cognitive outcome and treatment dose variables. **Abbreviations:** OTS=One-touch stockings of Cambridge (CANTAB).

**Supplementary Table S7.** Tertiary individual measures.

| | | Treatment completion | | | One-month follow-up | | |
|---|---|---|---|---|---|---|---|
| | Group | Treatment effect | 95% CI | *p*-value (adj.) | Treatment effect | 95% CI | *p*-value (adj.) |
| **Cognition** | | | | | | | |
| *Processing speed* | | | | | | | |
| RBANS Coding | H-CT | 0.16 | -0.14; 0.45 | 0.29 | 0.21 | -0.17; 0.59 | 0.28 |
| | TAU | - | - | - | - | - | - |
| Trail Making Test A | H-CT | 0.12 | -0.30; 0.53 | 0.58 | -0.21 | -0.76; 0.34 | 0.44 |
| | TAU | - | - | - | - | - | - |
| *Attention* | | | | | | | |
| RBANS Digit Span | H-CT | 0.50 | 0.06; 0.93 | **0.03** (0.20) | 0.29 | -0.37; 0.95 | 0.38 |
| | TAU | - | - | - | - | - | - |
| RVP accuracy | H-CT | 0.23 | -0.12; 0.57 | 0.20 | 0.38 | -0.001; 0.76 | 0.05 |
| | TAU | - | - | - | - | - | - |
| RVP mean latency | H-CT | 0.19 | -0.39; 0.76 | 0.52 | 0.26 | -0.12; 0.65 | 0.18 |
| | TAU | - | - | - | - | - | - |
| RVP false alarms | H-CT | -0.77 | -1.47; -0.08 | **0.03** (0.20) | -0.55 | -1.16; 0.06 | 0.08 |
| | TAU | - | - | - | - | - | - |
| RVP probability of hit | H-CT | 0.27 | -0.12; 0.67 | 0.17 | 0.42 | 0.01; 0.83 | **0.04** (0.21) |
| | TAU | - | - | - | - | - | - |
| *Working memory* | | | | | | | |
| WAIS-III Letter-number sequencing | H-CT | -0.17 | -0.72; 0.38 | 0.54 | 0.02 | -0.55; 0.60 | 0.93 |
| | TAU | - | - | - | - | - | - |
| SWM between errors | H-CT | 0.36 | -0.25; 0.96 | 0.24 | 0.10 | -0.45; 0.64 | 0.73 |
| | TAU | - | - | - | - | - | - |
| SWM strategy | H-CT | 0.21 | -0.28; 0.69 | 0.40 | 0.12 | -0.37; 0.60 | 0.63 |
| | TAU | - | - | - | - | - | - |
| *Verbal learning and memory* | | | | | | | |
| RAVLT total recall list I-V | H-CT | -0.18 | -0.61; 0.24 | 0.39 | 0.01 | -0.53; 0.55 | 0.97 |
| | TAU | - | - | - | - | - | - |

| | | | | | | | |
|---|---|---|---|---|---|---|---|
| RAVLT immediate recall | H-CT | -0.12 | -0.56; 0.32 | 0.58 | 0.67 | 0.11; 1.23 | **0.02** (0.19) |
| | TAU | - | - | - | - | - | - |
| RAVLT delayed recall | H-CT | 0.08 | -0.41; 0.57 | 0.74 | 0.15 | -0.37; 0.68 | 0.56 |
| | TAU | - | - | - | - | - | - |
| *Executive functions* | | | | | | | |
| Trail Making Test B | H-CT | 0.18 | -0.30; 0.66 | 0.46 | 0.09 | -0.39; 0.56 | 0.72 |
| | TAU | - | - | - | - | - | - |
| Verbal fluency letter S | H-CT | 0.37 | -0.07; 0.81 | 0.10 | 0.23 | -0.20; 0.66 | 0.28 |
| | TAU | - | - | - | - | - | - |
| Verbal fluency letter D | H-CT | 0.40 | 0.03; 0.77 | **0.03** (0.20) | 0.26 | -0.08; 0.60 | 0.12 |
| | TAU | - | - | - | - | - | - |
| OTS problems solved on first choice | H-CT | 0.66 | 0.03; 1.29 | **0.04** (0.21) | 0.92 | 0.23; 1.61 | **0.01** (0.11) |
| | TAU | - | - | - | - | - | - |
| OTS mean latency to correct | H-CT | -0.03 | -0.58; 0.52 | 0.91 | 0.05 | -0.51; 0.60 | 0.86 |
| | TAU | - | - | - | - | - | - |
| WCST perseverative errors | H-CT | 0.25 | -0.24; 0.73 | 0.31 | 0.45 | -0.20; 1.10 | 0.17 |
| | TAU | - | - | - | - | - | - |
| *Facial expression recognition* | | | | | | | |
| ERT hit rate across emotions | H-CT | 0.52 | 0.21; 0.82 | **0.001 (0.03)** | 0.34 | -0.06; 0.74 | 0.09 |
| | TAU | - | - | - | - | - | - |
| ERT overall reaction time | H-CT | 0.37 | -0.09; 0.83 | 0.11 | -0.21 | -0.75; 0.33 | 0.45 |
| | TAU | - | - | - | - | - | - |
| **Questionnaires/ratings** | | | | | | | |
| WHOQoL-BREF | H-CT | 0.34 | 0.01; 0.68 | **0.04** (0.21) | 0.28 | -0.12; 0.69 | 0.17 |
| | TAU | - | - | - | - | - | - |
| AQoL | H-CT | 0.55 | 0.20; 0.90 | **0.003 (0.04)** | 0.27 | -0.13; 0.66 | 0.18 |
| | TAU | - | - | - | - | - | - |
| COBRA | H-CT | 0.77 | 0.45; 1.08 | **<0.001 (0.006)** | 0.62 | 0.25; 1.00 | **0.002 (0.03)** |
| | TAU | - | - | - | - | - | - |
| FAST | H-CT | - | - | - | 0.17 | -0.17; 0.51 | 0.33 |
| | TAU | - | - | - | - | - | - |

| | | | | | | | |
|---|---|---|---|---|---|---|---|
| WSAS | H-CT | 0.29 | -0.05; 0.63 | 0.09 | 0.39 | -0.01; 0.78 | 0.06 |
| | TAU | - | - | - | - | - | - |
| SDS | H-CT | 0.37 | -0.08; 0.81 | 0.11 | 0.28 | -0.14; 0.70 | 0.18 |
| | TAU | - | - | - | - | - | - |
| PSQI | H-CT | 0.03 | -0.40; 0.46 | 0.89 | -0.09 | -0.51; 0.32 | 0.65 |
| | TAU | - | - | - | - | - | - |

**Note:** Table shows results from linear mixed models comparing H-CT and TAU from baseline to treatment completion and 1-month follow-up. **Sample sizes:** Baseline: 33 H-CT, 29 TAU. Treatment completion: 26 H-CT, 29 TAU. One-month follow-up: 26 H-CT, 29 TAU. **Missing data:** RBANS Coding: One participant at baseline (TAU). WCST perseverative errors: Three participants at baseline (one H-CT, two TAU), three participants at treatment completion (one H-CT, two TAU), five participants at follow-up (one H-CT, four TAU). ERT hit rate across emotions: four participants at baseline (one H-CT, three TAU), three participants at follow-up (one H-CT, two TAU). ERT overall reaction time: four participants at baseline (one H-CT, three TAU), three participants at follow-up (one H-CT, two TAU). WHOQoL-BREF: Three at baseline (two H-CT, one TAU), two at follow-up (one H-CT, one TAU). AQoL: Four at baseline (three H-CT, one TAU), two at follow-up (one H-CT, one TAU). COBRA: Two at baseline (one H-CT, one TAU), two at follow-up (one H-CT, one TAU). WSAS=Three at baseline (two H-CT, one TAU), two at follow-up (one H-CT, one TAU). SDS: Five at baseline (three H-CT, two TAU), two at follow-up (one H-CT, one TAU). PSQI: Three at baseline (two H-CT, one TAU), two at follow-up (one H-CT, one TAU). **Abbreviations:** Adj.=Adjusted; AQoL=Assessment of Quality of Life; CI=Confidence interval; COBRA=Cognitive Complaints in Bipolar Disorder Rating Assessment; ERT=Emotion Recognition Task; FAST=Functional Assessment Short Test; H-CT=Hypoxia with cognitive training; OTS=One-touch Stockings of Cambridge; PSQI=Pittsburgh Sleep Quality Index; RAVLT=Rey Auditory Verbal Learning Test; RBANS=Repeatable Battery for the Assessment of Neuropsychological Status; RVP=Rapid Visual Processing; TAU=Treatment as usual; SDS=Sheehan Disability Scale. SWM=Spatial Working Memory; WAIS=Wechsler Adult Intelligence Scale; WCST=Wisconsin Card Sorting Test; WHOQoL-BREF=World Health Organization Quality of Life; WSAS=Work and Social Adjustment Scale.

**Supplementary Table S8.** Change from before to after hypoxia with cognitive training treatment in primary, secondary, and tertiary outcomes in treatment as usual group participants who completed the intervention.

| | | **Treatment as usual participants who completed hypoxia cognitive training (*n*=18)** | | |
|---|---|---|---|---|
| | Timepoint | Treatment effect | 95% CI | *p*-value |
| **Primary outcome** | | | | |
| Speed of complex cognitive processing | End of treatment | 0.23 | -0.06; 0.52 | 0.11 |
| | Before treatment | - | - | - |
| **Secondary cognitive outcome** | | | | |
| OTS Mean choices to correct | End of treatment | 0.73 | -0.25; 1.71 | 0.13 |
| | Before treatment | - | - | - |
| **Secondary functioning outcome** | | | | |
| CAVIR global composite | End of treatment | 0.45 | 0.08; 0.83 | **0.02** |
| | Before treatment | - | - | - |
| **Tertiary outcomes** | | | | |
| **Cognitive domains** | | | | |
| Processing speed | End of treatment | 0.19 | -0.12; 0.50 | 0.20 |
| | Before treatment | - | - | - |
| Attention | End of treatment | 0.20 | -0.12; 0.52 | 0.20 |
| | Before treatment | - | - | - |
| Working memory | End of treatment | 0.35 | 0.03; 0.66 | **0.03** |
| | Before treatment | - | - | - |
| Verbal learning and memory | End of treatment | 0.60 | 0.13; 1.08 | **0.02** |
| | Before treatment | - | - | - |
| Executive functions | End of treatment | 0.16 | -0.10; 0.42 | 0.21 |
| | Before treatment | - | - | - |
| Global cognition | End of treatment | 0.30 | 0.07; 0.53 | **0.01** |
| | Before treatment | - | - | - |
| Facial emotion recognition | End of treatment | 0.17 | -0.06; 0.40 | 0.14 |
| | Before treatment | - | - | - |
| **Self-report/rating domains** | | | | |
| Quality of life | End of treatment | 0.18 | -0.25; 0.61 | 0.40 |
| | Before treatment | - | - | - |
| Subjective cognition | End of treatment | 0.37 | -0.05; 0.80 | 0.08 |
| | Before treatment | - | - | - |
| Functioning | End of treatment | -0.06 | -0.35; 0.23 | 0.67 |
| | Before treatment | - | - | - |

| | | | | |
|---|---|---|---|---|
| Sleep quality | End of treatment | 0.05 | -0.37; 0.46 | 0.81 |
| | Before treatment | - | - | - |

**Note:** Table shows results from linear mixed models investigating change in primary, secondary, and tertiary outcomes from before to end of hypoxia with cognitive training treatment for treatment as usual participants who subsequently completed the intervention and the exploratory end-of-treatment outcome assessment. **Missing data:** Facial emotion recognition: One participant before treatment. **Abbreviations:** CAVIR=Cognitive Assessment in Virtual Reality; CI=Confidence interval; OTS=One-touch Stockings of Cambridge (CANTAB).

**Supplementary Table S9.** Treatment group comparisons of average [$^{11}$C]UCB-J synaptic vesicle glycoprotein 2A (SV2A) binding as a readout for presynaptic density at treatment completion.

| | **H-CT** | **TAU** | **Effect size** | ***p*-value** |
|---|---|---|---|---|
| *N* | 13 | 15 | | |
| Hippocampus | 2.68 (0.34) | 2.92 (0.51) | 0.07 | 0.16 |
| Frontal cortex | 3.68 (0.47) | 3.90 (0.34) | 0.07 | 0.35 |
| Insula | 3.48 (0.48) | 3.77 (0.41) | 0.10 | 0.18 |
| Amygdala | 3.48 (0.46) | 3.73 (0.75) | 0.04 | 0.19 |
| Nucleus accumbens | 4.56 (0.48) | 5.00 (0.66) | 0.14 | **0.04** |
| Caudate nucleus | 4.01 (0.51) | 4.32 (0.55) | 0.08 | 0.19 |
| Putamen | 4.67 (0.62) | 4.99 (0.63) | 0.07 | 0.17 |
| Thalamus | 3.39 (0.43) | 3.63 (0.46) | 0.07 | 0.34 |
| Striatum | 4.34 (0.55) | 4.65 (0.57) | 0.08 | 0.17 |
| Neocortex | 3.65 (0.44) | 3.85 (0.35) | 0.07 | 0.35 |
| Anterior cingulate cortex | 3.88 (0.60) | 4.23 (0.52) | 0.10 | 0.19 |
| Posterior cingulate cortex | 4.08 (0.66) | 4.38 (0.47) | 0.07 | 0.31 |
| Gyrus rectus | 3.60 (0.66) | 4.03 (0.80) | 0.09 | 0.11 |
| Orbitofrontal gyri | 3.51 (0.45) | 3.78 (0.40) | 0.10 | 0.22 |
| Inferior frontal gyrus | 3.87 (0.49) | 4.10 (0.34) | 0.08 | 0.28 |
| Superior frontal gyrus | 3.64 (0.50) | 3.84 (0.35) | 0.06 | 0.44 |
| Middle frontal gyrus | 3.78 (0.52) | 3.94 (0.32) | 0.04 | 0.61 |
| Precentral gyrus | 3.49 (0.49) | 3.63 (0.27) | 0.04 | 0.70 |
| Postcentral gyrus | 3.47 (0.48) | 3.63 (0.33) | 0.05 | 0.62 |
| Superior parietal gyrus | 3.71 (0.43) | 3.82 (0.35) | 0.02 | 0.83 |
| Inferolateral remainder of parietal lobe | 3.87 (0.50) | 4.03 (0.36) | 0.04 | 0.58 |
| Anterior temporal lobe (medial) | 3.24 (0.45) | 3.54 (0.56) | 0.08 | 0.14 |
| Anterior temporal lobe (lateral) | 3.80 (0.45) | 4.07 (0.56) | 0.07 | 0.20 |
| Posterior temporal lobe | 3.72 (0.47) | 3.93 (0.46) | 0.05 | 0.36 |

| | | | | |
|---|---|---|---|---|
| Parahippocampal and ambient gyri | 2.86 (0.43) | 3.15 (0.48) | 0.10 | 0.13 |
| Superior temporal gyrus | 3.76 (0.51) | 4.06 (0.37) | 0.12 | 0.20 |
| Middle and inferior temporal gyri | 3.91 (0.46) | 4.18 (0.50) | 0.08 | 0.17 |
| Fusiform gyrus | 3.53 (0.52) | 3.82 (0.58) | 0.07 | 0.20 |
| Lingual gyrus | 3.71 (0.50) | 3.94 (0.40) | 0.07 | 0.31 |
| Cuneus | 3.67 (0.45) | 3.91 (0.41) | 0.08 | 0.31 |
| Lateral remainder of occipital lobe | 3.46 (0.34) | 3.65 (0.42) | 0.06 | 0.35 |

**Note:** Values are shown in raw means and standard deviations. All regions are bilateral. Effect sizes were computed as sizes were computed as partial eta-squared, where 0.01 are interpreted as small, 0.06 as medium, and >0.14 as large effects. **Abbreviations:** H-CT=Hypoxia with cognitive training; TAU=Treatment as usual.

**Supplementary Table S10.** Cluster maxima table.

| | | | | | MNI coordinates | | |
|---|---|---|---|---|---|---|---|
| | BA | *p* | Cluster size (no. Voxels) | Peak Z | X | Y | Z |
| dPFC VOI | | | | | | | |
| 1>0-back | | | | | | | |
| Right precentral gyrus | 6 | **0.002** | 262 | 4.66 | 40 | -4 | 58 |
| Left precentral gyrus | 6 | **0.03** | 153 | 4.16 | -38 | 8 | 28 |
| Whole brain analysis | | | | | | | |
| 2>0-back | | | | | | | |
| Right lateral occipital cortex, superior division | 7 | **0.005** | 301 | 3.84 | 18 | -78 | 52 |
| 1>0-back | | | | | | | |
| Right precentral gyrus | 6 | **<0.001** | 1941 | 4.66 | 40 | -4 | 58 |
| Left postcentral gyrus | 1 | **<0.001** | 768 | 4.07 | -52 | -16 | 36 |
| Right lateral occipital cortex, inferior division | 18 | **<0.001** | 442 | 3.56 | 42 | -86 | -10 |

**Note:** Table shows peak cluster activation in regions showing differential change in neural response during spatial working memory N-back task over time in hypoxia with cognitive training compared to treatment as usual groups. **Abbreviations:** BA=Brodmann area; dPFC=Dorsal prefrontal cortex; MNI=Montreal Neurological Institute; VOI=Volume of interest.

**Supplementary Table S11.** Correlation matrix of change in neural response and change in neuropsychological outcomes in the hypoxia with cognitive training group.

| | **Completed hypoxia with cognitive training and fMRI (*n*=19)** | | | | | | | | | | | |
|---|---|---|---|---|---|---|---|---|---|---|---|---|
| | **One-month follow-up** | | | | | | | | | | | |
| | ΔdPFC right precentral gyrus 1>0-back | | ΔdPFC left precentral gyrus 1>0-back | | ΔWhole brain right precentral gyrus 1>0-back | | ΔWhole brain left postcentral gyrus 1>0-back | | ΔWhole brain right lateral occipital cortex 1>0-back | | ΔWhole brain right lateral occipital cortex 2>0-back | |
| | *r* | *p*-value | *r* | *p*-value | *r* | *p*-value | *r* | *p*-value | *r* | *p*-value | *r* | *p*-value |
| Δ*d*' | -0.43 | 0.08 | -0.37 | 0.13 | -0.15 | 0.54 | -0.10 | 0.68 | -0.40 | 0.10 | -0.46 | 0.06 |
| ΔOTS 'mean choices to correct' | 0.35 | 0.14 | 0.04 | 0.87 | -0.14 | 0.57 | 0.14 | 0.56 | 0.10 | 0.67 | -0.03 | 0.92 |
| ΔExecutive functions | 0.03 | 0.89 | 0.09 | 0.72 | 0.10 | 0.67 | 0.13 | 0.61 | 0.10 | 0.69 | 0.11 | 0.66 |
| ΔWorking memory | -0.51 | **0.03** | 0.15 | 0.54 | 0.18 | 0.46 | 0.01 | 0.97 | 0.01 | 0.98 | 0.13 | 0.59 |
| ΔFunctioning | -0.17 | 0.48 | 0.002 | 0.995 | 0.33 | 0.17 | -0.26 | 0.28 | -0.08 | 0.74 | 0.10 | 0.67 |
| ΔSubjective cognition | -0.36 | 0.14 | 0.23 | 0.36 | 0.23 | 0.36 | -0.10 | 0.70 | 0.10 | 0.69 | 0.11 | 0.67 |

**Note:** Table shows correlation matrix of change-scores in relevant behavioral and self-reported measures and dPFC and whole-brain clusters showing de-activation over time in H-CT group from baseline to one-month follow-up. Correlations with *d*' made with 1-back *d'* for 1>0-back contrasts and combined 1-back and 2-back *d*' for 2>0-back contrasts. **Abbreviations:** CI=Confidence interval; *d*'=Discriminability index; dPFC=Dorsal prefrontal cortex; fMRI=Functional magnetic resonance imaging; H-CT=Hypoxia with cognitive training; OTS=One-touch stockings of Cambridge.

**Supplementary Table S12.** Exploratory, peripheral neuroplasticity marker outcomes.

| | | **Day 19** | | |
|---|---|---|---|---|
| | Group | Treatment effect | 95% CI | *p*-value |
| EPO | H-CT | -0.49 | -1.14; 0.16 | 0.14 |
| | TAU | - | - | - |
| BDNF | H-CT | -0.31 | -0.68; 0.06 | 0.10 |
| | TAU | - | - | - |
| VEGF | H-CT | 0.10 | -0.17; 0.37 | 0.44 |
| | TAU | - | - | - |

**Note:** Table shows results from linear mixed models comparing H-CT and TAU from day 1 to day 19 on serum EPO, BDNF, and VEGF. **Blood test sample sizes:** Day 1: 29 H-CT, 27 TAU. Day 19: 25 H-CT, 27 TAU. **Abbreviations:** BDNF=Brain-derived neurotrophic factor; EPO=Erythropoietin; H-CT=Hypoxia with cognitive training; SD=Standard deviation; TAU=Treatment as usual; VEGF=Vascular endothelial growth factor.

### Supplementary Figures

**Supplementary Figure S1.** Flow chart for the subsample of participants included in the PET imaging analysis.

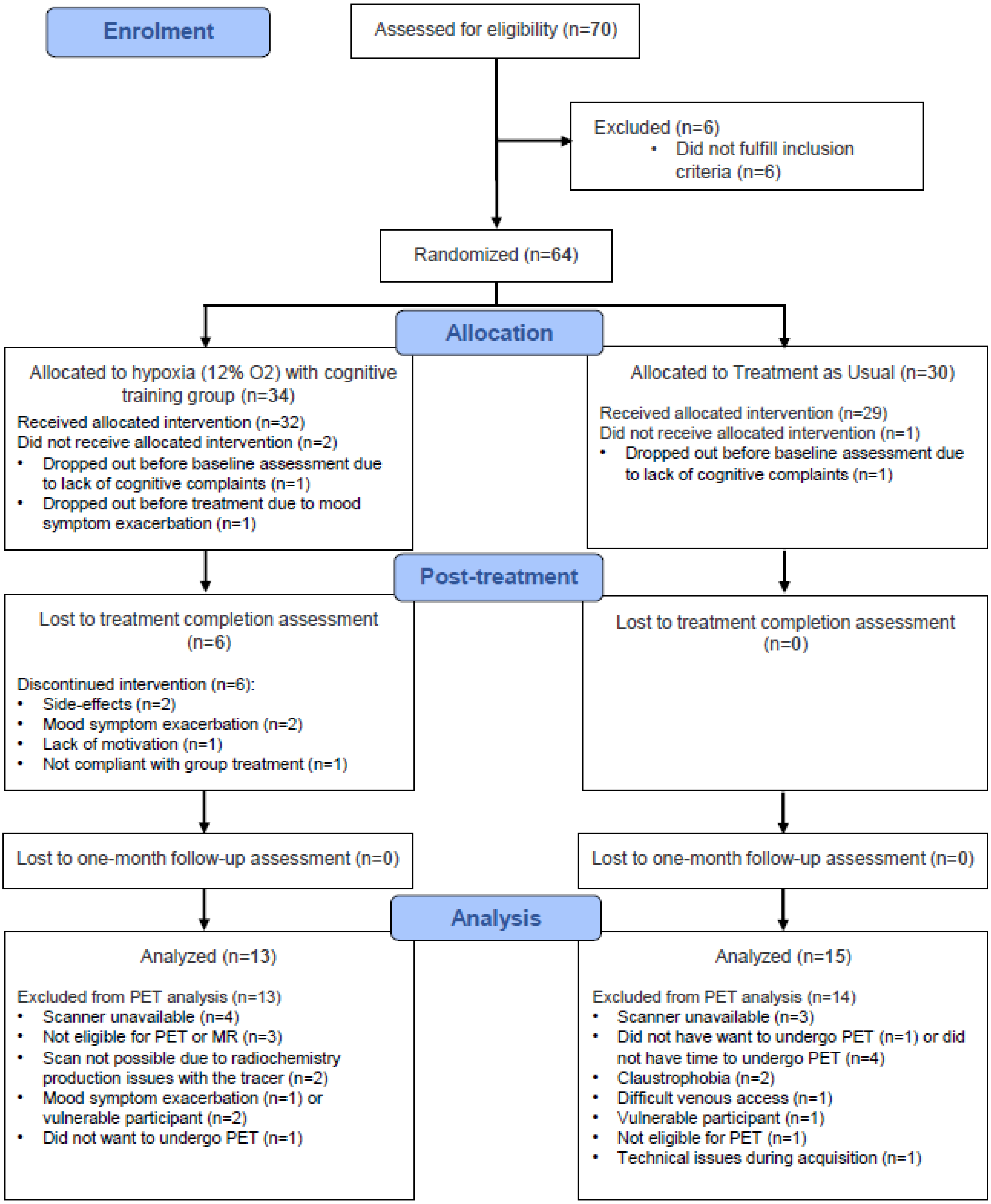

**Supplementary Figure S2.** Hematological parameters changes across hypoxia with cognitive training, treatment as usual, and healthy participant hypoxia and normoxia groups.

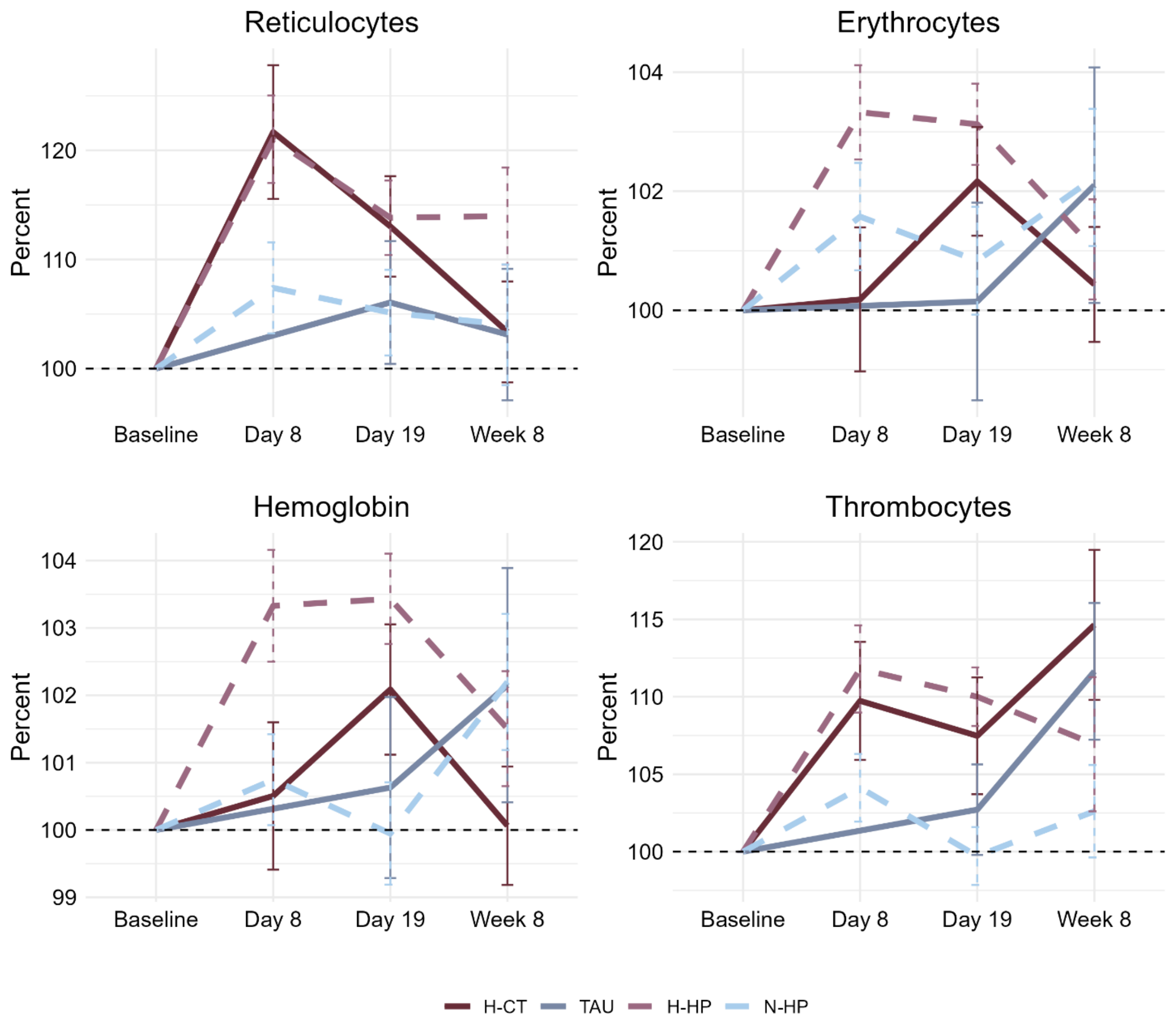

**Note:** Values are shown in percent change from baseline (mean for each group for each time point). Error bars show the standard error. **Abbreviations:** H-CT=Hypoxia with cognitive training; TAU=Treatment as usual; H-HP=Hypoxia (healthy individuals); N-HP=Normoxia (healthy individuals).

**Supplementary Figure S3.** Mean percent signal change in dPFC clusters showing differential change in neural activity over time during low-load working memory in hypoxia with cognitive training vs. treatment as usual groups.

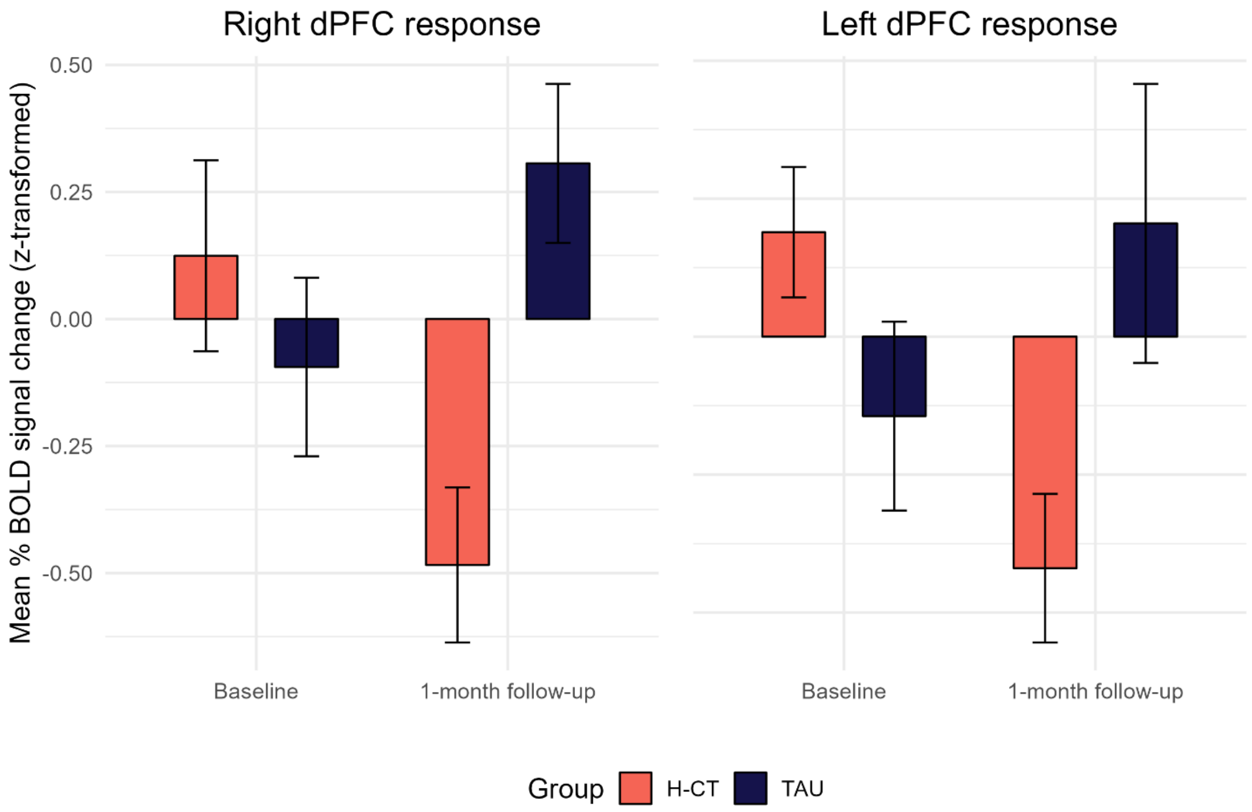

**Note:** Bar heights represent mean values per group and time point. Error bars represent standard errors of the mean. **Abbreviations:** BOLD=Blood-oxygen-level-dependent; dPFC=Dorsal prefrontal cortex; H-CT=Hypoxia with cognitive training; HC=Healthy controls; TAU=Treatment as usual.
