## Supplementary material for "Moderate hypoxia and cognitive training for cognitive impairment in mood disorders: a randomized controlled trial": CONSORT checklist

|  | Section/topic | No | CONSORT 2025 checklist item description | Reported on page no. |
| --- | --- | --- | --- | --- |
|  | **Title and abstract** | | |  |
|  | Title and structured abstract | 1a | Identification as a randomised trial | Title |
|  |  | 1b | Structured summary of the trial design, methods, results, and conclusions | Abstract |
|  | **Open science** | | |  |
|  | Trial registration | 2 | Name of trial registry, identifying number (with URL) and date of registration | p.1 |
|  | Protocol and statistical analysis plan | 3 | Where the trial protocol and statistical analysis plan can be accessed | p.1 |
|  | Data sharing | 4 | Where and how the individual de-identified participant data (including data dictionary), statistical code and any other materials can be accessed | p.21 |
|  | Funding and conflicts of interest | 5a | Sources of funding and other support (eg, supply of drugs), and role of funders in the design, conduct, analysis and reporting of the trial | p.21 |
|  |  | 5b | Financial and other conflicts of interest of the manuscript authors | p.22 |
|  | **Introduction** | | |  |
|  | Background and rationale | 6 | Scientific background and rationale | pp.3-5 |
|  | Objectives | 7 | Specific objectives related to benefits and harms | pp.4-5 |
|  | **Methods** | | |  |
|  | Patient and public involvement | 8 | Details of patient or public involvement in the design, conduct and reporting of the trial | p.5 |
|  | Trial design | 9 | Description of trial design including type of trial (eg, parallel group, crossover), allocation ratio, and framework (eg, superiority, equivalence, non-inferiority, exploratory) | p.5 |
|  | Changes to trial protocol | 10 | Important changes to the trial after it commenced including any outcomes or analyses that were not prespecified, with reason | p.6 |
|  | Trial setting | 11 | Settings (eg, community, hospital) and locations (eg, countries, sites) where the trial was conducted | p.5 |
|  | Eligibility criteria | 12a | Eligibility criteria for participants | p.6, p.S3 |
|  |  | 12b | If applicable, eligibility criteria for sites and for individuals delivering the interventions (eg, surgeons, physiotherapists) | N/A (single-site study) |
|  | Intervention and comparator | 13 | Intervention and comparator with sufficient details to allow replication. If relevant, where additional materials describing the intervention and comparator (eg, intervention manual) can be accessed | pp.7-8, pp.S4-S6 |
|  | Outcomes | 14 | Prespecified primary and secondary outcomes, including the specific measurement variable (eg, systolic blood pressure), analysis metric (eg, change from baseline, final value, time to event), method of aggregation (eg, median, proportion), and time point for each outcome | pp.8-9 |
|  | Harms | 15 | How harms were defined and assessed (eg, systematically, non-systematically) | pp.7-8, p.S6 |
|  | Sample size | 16a | How sample size was determined, including all assumptions supporting the sample size calculation | pp.11-12, pp.S8-S9 |
|  |  | 16b | Explanation of any interim analyses and stopping guidelines | N/A (no interim analyses or stopping guidelines) |
|  | Randomisation: |  |  |  |
|  | Sequence generation | 17a | Who generated the random allocation sequence and the method used | p.6 |
|  |  | 17b | Type of randomisation and details of any restriction (eg, stratification, blocking and block size) | p.6 |
|  | Allocation concealment mechanism | 18 | Mechanism used to implement the random allocation sequence (eg, central computer/telephone; sequentially numbered, opaque, sealed containers), describing any steps to conceal the sequence until interventions were assigned | p.6 |
|  | Implementation | 19 | Whether the personnel who enrolled and those who assigned participants to the interventions had access to the random allocation sequence | p.6 |
|  | Blinding | 20a | Who was blinded after assignment to interventions (eg, participants, care providers, outcome assessors, data analysts) | p.6 |
|  |  | 20b | If blinded, how blinding was achieved and description of the similarity of interventions | N/A (participants not blinded) |
|  | Statistical methods | 21a | Statistical methods used to compare groups for primary and secondary outcomes, including harms | p.11-12, p.S6 |
|  |  | 21b | Definition of who is included in each analysis (eg, all randomised participants), and in which group | pp.12-13, Figure 1, Figure S1 |
|  |  | 21c | How missing data were handled in the analysis | pp.11-13, p.S6 |
|  |  | 21d | Methods for any additional analyses (eg, subgroup and sensitivity analyses), distinguishing prespecified from post hoc | pp.10-11, pp.S7-S15 |
|  | **Results** | | |  |
|  | Participant flow, including flow diagram | 22a | For each group, the numbers of participants who were randomly assigned, received intended intervention, and were analysed for the primary outcome | pp.12-13,  Figure 1 |
|  |  | 22b | For each group, losses and exclusions after randomisation, together with reasons | pp.12-13, Figure 1 |
|  | Recruitment | 23a | Dates defining the periods of recruitment and follow-up for outcomes of benefits and harms | p.12 |
|  |  | 23b | If relevant, why the trial ended or was stopped | N/A (did not end prematurely) |
|  | Intervention and comparator delivery | 24a | Intervention and comparator as they were actually administered (eg, where appropriate, who delivered the intervention/comparator, how participants adhered, whether they were delivered as intended (fidelity)) | pp.13-14, p.S16, Table S2 |
|  |  | 24b | Concomitant care received during the trial for each group | N/A (no concominant care) |
|  | Baseline data | 25 | A table showing baseline demographic and clinical characteristics for each group | Table 1, Table S1 |
|  | Numbers analysed,  outcomes and estimation | 26 | For each primary and secondary outcome, by group:  ● the number of participants included in the analysis  ● the number of participants with available data at the outcome time point  ● result for each group, and the estimated effect size and its precision (such as 95% confidence interval)  ● for binary outcomes, presentation of both absolute and relative effect size | p.14,  Tables 2-3 |
|  | Harms | 27 | All harms or unintended events in each group | pp.13-14, pp.S17-S18, Tables S3-S4 |
|  | Ancillary analyses | 28 | Any other analyses performed, including subgroup and sensitivity analyses, distinguishing pre-specified from post hoc | pp.15-17, pp.S18-S19, Tables S5-S12 |
|  | **Discussion** | | |  |
|  | Interpretation | 29 | Interpretation consistent with results, balancing benefits and harms, and considering other relevant evidence | pp.17-21 |
|  | Limitations | 30 | Trial limitations, addressing sources of potential bias, imprecision, generalisability, and, if relevant, multiplicity of analyses | pp.20-21 |

Citation: Hopewell S, Chan AW, Collins GS, Hróbjartsson A, Moher D, Schulz KF, et al. CONSORT 2025 Statement: updated guideline for reporting randomised trials. BMJ. 2025; 388:e081123. <https://dx.doi.org/10.1136/bmj-2024-081123>
© 2025 Hopewell et al. This is an Open Access article distributed under the terms of the Creative Commons Attribution License (<https://creativecommons.org/licenses/by/4.0/>), which permits unrestricted use, distribution, and reproduction in any medium, provided the original work is properly cited.

*We strongly recommend reading this statement in conjunction with the CONSORT 2025 Explanation and Elaboration and/or the CONSORT 2025 Expanded Checklist for important clarifications on all the items. We also recommend reading relevant CONSORT extensions. See [www.consort-spirit.org](http://www.consort-spirit.org).
